# Clinical Impact of Pharmacogenetic Risk Variants in a Large Chinese Cohort

**DOI:** 10.1101/2024.10.15.24315139

**Authors:** Chun-Yu Wei, Ming-Shien Wen, Chih-Kuang Cheng, Yi-Jing Sheen, Tsung-Chieh Yao, Sing-Lian Lee, Jer-Yuarn Wu, Ming-Fang Tsai, Ling-Hui Li, Chun-Houh Chen, Cathy S.-J. Fann, Hsin-Chou Yang, Yen-Tsung Huang, Hung-Hsin Chen, Yi-Min Liu, Erh-Chan Yeh, Yu-Ching Peng, Shuu-Jiun Wang, Shih-Pin Chen, Ming-Tsun Tsai, Teh-Ia Huo, Chien-Wei Su, Der-Cherng Tarng, Chin-Chou Huang, Jong-Ling Fuh, Keng-Hsin Lan, Yo-Tsen Liu, Ching-Liang Lu, Yi-Chung Lee, Yi-Hsiang Huang, Chung-Pin Li, Yen-Feng Wang, Yu-Cheng Hsieh, Yi-Ming Chen, Tzu-Hung Hsiao, Ching-Heng Lin, Yen-Ju Chen, I-Chieh Chen, Chien-Lin Mao, Shu-Jung Chang, Yen-Lin Chang, Yi-Ju Liao, Chih-Hung Lai, Wei-Ju Lee, Hsin Tung, Ting-Ting Yen, Hsin-Chien Yen, Jer-Hwa Chang, Chun-Yao Huang, Lung Chan, Yung-Wei Lin, Bu-Yuan Hsiao, Chaur-Jong Hu, Yung-Kuo Lin, Yung-Feng Lin, Tung-Cheng Chang, Deng-Chyang Wu, Jung-Yu Kan, Chung-Yao Hsu, Szu-Chia Chen, Ching-Chia Li, Chung-Feng Huang, Chau-Chyun Sheu, Lii-Jia Yang, Chung-Hwan Chen, Kuan-Mao Chen, Shu-Min Chang, Min-Shiuan Liou, Shi-Ping Wang, Kuan-Ting Lin, Hui-Ping Chuang, Ying-Ju Chen, Joey Sin, Ying-Ting Chen, Chiung-Chih Chang, Chang-Fu Kuo, Jing-Chi Lin, Ho-Chang Kuo, Tien-Min Chan, Chao-Wei Lee, Jenn-Haung Lai, Shue-Fen Luo, Hao-Tsai Cheng, Lian-Yu Lin, Li-Chun Chang, Chia-Ti Tsai, Hsien-Li Kao, Jian-Jyun Yu, Jiann-Shing Jeng, Min-Chin Chiu, Tzu-Chan Hong, Shun-Fa Yang, Hsueh-Ju Lu, Sheng-Chiang Su, Pau-Ling Chu, Peng-Fei Li, Chia-Lin Tsai, Chia-Kuang Tsai, Shih-En Tang, Chien-Ming Lin, Yung-Fu Wu, Chih-Yang Huang, Shinn-Zong Ling, Chun-Chun Chang, Tzu-Kai Lin, Sheng-Mou Hsiao, Chih-Hung Chang, Chih-Dao Chen, Gwo-Chin Ma, Ting-Yu Chang, Juey-Jen Hwang, Chien-Lin Lu, Kuo-Jang Kao, Chen-Fang Hung, Shiou-Sheng Chen, Po-Yueh Chen, Ko-Chung Tsui, Chien-Hsiun Chen, Chih-Cheng Chien, Han-Sun Chiang, Yen-Ling Chiu, Hsiang-Cheng Chen, Pui-Yan Kwok

**Affiliations:** Institute of Biomedical Sciences, Academia Sinica, Taipei, Taiwan; Core Laboratory of Neoantigen Analysis for Personalized Cancer Vaccine, Office of R&D, Taipei Medical University, Taipei, Taiwan; Division of Cardiology, Chang Gung Memorial Hospital, Linkou Branch, Taoyuan, Taiwan; School of Medicine, Chang Gung University, Taoyuan, Taiwan; Chang Gung Memorial Hospital, Linkou Branch, Taoyuan, Taiwan; Division of Endocrinology and Metabolism, Department of Internal Medicine, Taichung Veterans General Hospital, Taiwan; Department of Medicine, School of Medicine, National Yang Ming Chiao Tung University, Taipei, Taiwan; Department of Post-Baccalaureate Medicine, College of Medicine, National Chung Hsing University, Taichung, Taiwan; Division of Allergy, Asthma, and Rheumatology, Department of Pediatrics, Chang Gung Memorial Hospital, Taoyuan, Taiwan; Department of Internal Medicine, Koo Foundation Sun Yat-Sen Cancer Center, Taipei, Taiwan; Institute of Statistical Science, Academia Sinica, Taipei, Taiwan; Biomedical Translation Research Center, Academia Sinica, Taipei, Taiwan; Bioinformatics Program, Taiwan International Graduate Program, Academia Sinica, Taipei, Taiwan; Department of Mathematics, Institute of Epidemiology and Preventive Medicine, National Taiwan University, Taiwan; Department of Pathology and Laboratory Medicine, Taipei Veterans General Hospital, Taipei, Taiwan; Department of Neurology, Neurological Institute, Taipei Veterans General Hospital, Taipei, Taiwan; College of Medicine, National Yang Ming Chiao Tung University, Taipei, Taiwan; Brain Research Center, National Yang Ming Chiao Tung University, Taipei, Taiwan; Division of Translational Research, Department of Medical Research, Taipei Veterans General Hospital, Taipei, Taiwan; Institute of Clinical Medicine, National Yang Ming Chiao Tung University, Taipei, Taiwan; Division of Nephrology, Department of Medicine, Taipei Veterans General Hospital, Taipei, Taiwan; Division of Basic Research, Department of Medical Research, Taipei Veterans General Hospital, Taipei, Taiwan; Institute of Pharmacology, National Yang Ming Chiao Tung University, Taipei, Taiwan; School of Medicine, National Yang Ming Chiao Tung University, Taipei, Taiwan; Division of General Medicine, Department of Medicine, Taipei Veterans General Hospital, Taipei, Taiwan; Faculty of Medicine, School of Medicine, National Yang Ming Chiao Tung University, Taipei, Taiwan; Institute of Clinical Medicine, School of Medicine, National Yang Ming Chiao Tung University, Taipei, Taiwan; Division of Cardiology, Department of Medicine, Taipei Veterans General Hospital, Taipei, Taiwan; Brain Research Center & School of Medicine, National Yang Ming Chiao Tung University, Taipei, Taiwan; Division of Gastroenterology and Hepatology, Department of Medicine, Taipei Veterans General Hospital, Taipei, Taiwan; Institute of Pharmacology, College of Medicine, National Yang Ming Chiao Tung University, Taipei, Taiwan; Division of Epilepsy, Neurological Institute, Taipei Veterans General Hospital, Taipei, Taiwan; Institute of Brain Science, National Yang Ming Chiao Tung University, Taipei, Taiwan; Division of Gastroenterology, Taipei Veterans General Hospital, Taipei, Taiwan; Department of Neurology, National Yang Ming Chiao Tung University School of Medicine, Taipei, Taiwan; Brain Research Center, National Yang-Ming University, Taipei, Taiwan; Healthcare and Services Center, Taipei Veterans General Hospital, Taipei, Taiwan; Institute of Clinical Medicine, College of Medicine, National Yang Ming Chiao Tung University, Taipei, Taiwan; Division of Clinical Skills Training, Department of Medical Education, Taipei Veterans General Hospital, Taipei, Taiwan; School of Medicine, College of Medicine, National Yang Ming Chiao Tung University, Taipei, Taiwan; Therapeutic and Research Center of Pancreatic Cancer, Taipei Veterans General Hospital, Taipei, Taiwan; Department of Medical Research, Taichung Veterans General Hospital, Taichung, Taiwan; Department of Post-Baccalaureate Medicine, National Chung Hsing University, Taichung, Taiwan; Institute of Clinical Medicine, National Yang Ming Chiao Tung University, Taipei, Taiwan; Department of Medical Research, Taiwan; Department of Pharmacy, Taichung Veterans General Hospital, Taichung, Taiwan; Department of Medicine and Cardiovascular Center, Taichung Veterans General Hospital, Taichung, Taiwan; Neurological Institute, Taichung Veterans General Hospital, Taichung, Taiwan; Department of Otolaryngology, Taichung Veterans General Hospital, Taichung, Taiwan; Division of Pediatric Genetics and Metabolism, Children’s Medical Center, Taichung Veterans General Hospital, Taichung, Taiwan; School of Respiratory Therapy, College of Medicine, Taipei Medical University, Taipei, Taiwan; Division of Pulmonary Medicine, Department of Internal Medicine, Wan Fang Hospital, Taipei Medical University, Taipei, Taiwan; Division of Cardiology, Department of Internal Medicine, School of Medicine, College of Medicine, Taipei Medical University, Taipei, Taiwan; Division of Cardiology and Cardiovascular Research Center, Department of Internal Medicine, Taipei Medical University Hospital, Taipei, Taiwan; Taipei Heart Institute, Taipei Medical University, Taipei, Taiwan; Department of Neurology, Shuang Ho Hospital, Taipei Medical University, New Taipei City, Taiwan; Department of Neurology, School of Medicine, College of Medicine, Taipei Medical University, Taipei, Taiwan; Taipei Neuroscience Institute, Shuang Ho Hospital, Taipei Medical University, New Taipei City, Taiwan; Department of Urology, School of Medicine, College of Medicine and TMU Research Center of Urology and Kidney (TMU-RCUK), Taipei Medical University, Taipei, Taiwan; Department of Urology, Wan Fang Hospital, Taipei Medical University, Taipei, Taiwan; International Master/PhD Program in Medicine, College of Medicine, Taipei Medical University, Taipei, Taiwan; Division of Cardiovascular Medicine, Department of Internal Medicine, Wan Fang Hospital, Taipei Medical University, Taipei, Taiwan; Department of Medicine Research, Taipei Medical University Hospital, Taipei, Taiwan; Division of Colorectal Surgery, Department of Surgery, Shuang Ho Hospital, Taipei Medical University, New Taipei City, Taiwan; Division of General Surgery, Department of Surgery, School of Medicine, College of Medicine, Taipei Medical University, Taipei, Taiwan; Division of Gastroenterology, Department of Internal Medicine, Kaohsiung Medical University Hospital, Kaohsiung Medical University, Kaohsiung, Taiwan; Department of Medicine, Faculty of Medicine, College of Medicine, Kaohsiung Medical University, Kaohsiung, Taiwan; Department of Internal Medicine, Kaohsiung Medical University Gangshan Hospital, Kaohsiung, Taiwan; Division of Breast Oncology and Surgery, Department of Surgery, Kaohsiung Medical University Chung-Ho Memorial Hospital, Kaohsiung Medical University, Kaohsiung, Taiwan; Department of Post Baccalaureate Medicine, College of Medicine, Kaohsiung Medical University, Kaohsiung, Taiwan; Department of Surgery, Faculty of Medicine, College of Medicine, Kaohsiung Medical University, Kaohsiung, Taiwan; Sleep Medicine Center, Department of Neurology, Kaohsiung Medical University Hospital, Kaohsiung Medical University, Kaohsiung, Taiwan; Department of Neurology, College of Medicine, Kaohsiung Medical University, Kaohsiung, Taiwan; Division of Nephrology, Department of Internal Medicine, Kaohsiung Medical University Hospital, Kaohsiung Medical University, Kaohsiung, Taiwan; Department of Internal Medicine, Kaohsiung Municipal Siaogang Hospital, Kaohsiung Medical University, Kaohsiung, Taiwan; Faculty of Medicine, College of Medicine, Kaohsiung Medical University, Kaohsiung, Taiwan; Department of Urology, Kaohsiung Medical University Hospital, Kaohsiung Medical University, Kaohsiung, Taiwan; Department of Urology, Kaohsiung Medical University Gangshan Hospital, Kaohsiung, Taiwan; Hepatobiliary Division, Department of Internal Medicine, Kaohsiung Medical University Hospital, Kaohsiung Medical University, Kaohsiung, Taiwan; Faculty of Internal Medicine and Hepatitis Research Center, School of Medicine, College of Medicine, Kaohsiung Medical University, Kaohsiung, Taiwan; Division of Pulmonary and Critical Care Medicine, Department of Internal Medicine, Kaohsiung Medical University Hospital, Kaohsiung Medical University, Kaohsiung, Taiwan; Department of Internal Medicine, School of Medicine, College of Medicine, Kaohsiung Medical University, Kaohsiung, Taiwan; Department of Internal Medicine, Kaohsiung Municipal CiJin Hospital, Kaohsiung, Taiwan; Orthopaedic Research Center and Department of Orthopedics, College of Medicine, Kaohsiung Medical University, Kaohsiung, Taiwan; Regenerative Medicine and Cell Therapy Research Center, Kaohsiung Medical University, Kaohsiung, Taiwan; Department of Orthopedics, Kaohsiung Municipal Ta-Tung Hospital and Kaohsiung Medical University Hospital, Kaohsiung, Taiwan; Department of Neurology, Cognition and Aging Center, Kaohsiung Chang Gung Memorial Hospital, Kaohsiung, Taiwan; Institute for Translational Research in Biomedicine, Kaohsiung Chang Gung Memorial Hospital, Kaohsiung, Taiwan; School of Medicine, College of Medicine, National Sun Yat-sen University, Kaohsiung, Taiwan; Center for Artificial Intelligence in Medicine, Chang Gung Memorial Hospital, Taoyuan, Taiwan; Division of Rheumatology, Allergy and Immunology, Chang Gung Memorial Hospital, Taoyuan, Taiwan; Department of Internal Medicine, College of Medicine, Chang Gung University, Taoyuan, Taiwan; Division of Allergy Immunology and Rheumatology, Chang Gung Memorial Hospital, Chiayi, Taiwan; Pediatric Internal Medicine, Department of Internal Medicine, Kaohsiung Chang Gung Memorial Hospital, Taiwan; Chang Gung University College of Medicine, Kaohsiung, Taiwan; Division of General Surgery, Department of Surgery, Linkou Chang Gung Memorial Hospital, Taoyuan, Taiwan; Department of Rheumatology, Allergy and Immunology, Linkou Chang Gung Memorial Hospital, Taoyuan, Taiwan; Department of Gastroenterology and Hepatology, New Taipei Municipal TuCheng Hospital New Taipei City, Taiwan; Department of Gastroenterology and Hepatology, Linkou Chang Gung Memorial Hospital, Taoyuan, Taiwan; Graduate Institute of Clinical Medicine, College of Medicine, Chang Gung University, Taoyuan, Taiwan; Department of Internal Medicine, National Taiwan University Hospital, Taipei, Taiwan; Department of Internal Medicine, National Taiwan University College of Medicine, Taipei, Taiwan; Department of Internal Medicine, National Taiwan University Hospital, Yunlin Branch, Yunlin, Taiwan; Department of Neurology, National Taiwan University Hospital, Taipei, Taiwan; Department of Neurology, National Taiwan University College of Medicine, Taipei, Taiwan; Department of Internal Medicine, National Taiwan University Cancer Center, Taipei, Taiwan; Graduate Institute of Clinical Medicine, National Taiwan University College of Medicine, Taiwan; Institute of Medicine, Chung Shan Medical University, Taichung, Taiwan; Department of Medical Research, Chung Shan Medical University Hospital, Taichung, Taiwan; Division of Hematology and Oncology, Department of Internal Medicine, Chung Shan Medical University Hospital, Taichung, Taiwan; School of Medicine, Chung Shan Medical University, Taichung, Taiwan; Endocrinology and Metabolism, Tri-Service General Hospital, National Defense Medical Center, Taipei, Taiwan; Nephrology, Tri-Service General Hospital, National Defense Medical Center, Taipei, Taiwan; Pulmonary Medicine, Tri-Service General Hospital, National Defense Medical Center, Taipei, Taiwan; Pediatrics, Tri-Service General Hospital, National Defense Medical Center, Taipei, Taiwan; Department of Medical Research, Tri-Service General Hospital, National Defense Medical Center, Taipei, Taiwan; Cardiovascular and Mitochondria Related Disease Research Center, Hualien Tzu Chi Hospital, Buddhist Tzu Chi Medical Foundation, Hualien, Taiwan; Center of General Education, Buddhist Tzu Chi Medical Foundation, Tzu Chi University of Science and Technology, Hualien, Taiwan; Bioinnovation Center, Buddhist Tzu Chi Medical Foundation, Hualien, Taiwan; Department of Neurosurgery, Hualien Tzu Chi Hospital, Buddhist Tzu Chi Medical Foundation, Hualien, Taiwan; Department of Laboratory Medicine, Hualien Tzu Chi Hospital, Buddhist Tzu Chi Medical Foundation, Hualien, Taiwan; Department of Laboratory Medicine and Biotechnology, Tzu Chi University, Hualien, Taiwan; Department of Dermatology, Hualien Tzu Chi Hospital, Buddhist Tzu Chi Medical Foundation, Hualien, Taiwan; School of Medicine, Tzu Chi University, Hualien, Taiwan; Department of Obstetrics and Gynecology, Far Eastern Memorial Hospital, New Taipei, Taiwan; Graduate School of Biotechnology and Bioengineering, Yuan Ze University, Taoyuan, Taiwan; Department of Obstetrics and Gynecology, National Taiwan University Hospital, Taipei, Taiwan; Department of Orthopedic Surgery, Far Eastern Memorial Hospital, New Taipei City, Taiwan; Department of Family Medicine, Far Eastern Memorial Hospital, New Taipei City, Taiwan; Department of Genomic Medicine and Center for Medical Genetics, Changhua Christian Hospital, Changhua, Taiwan; Department of Cardiology, Fu Jen Catholic University Hospital, Fu Jen Catholic University, New Taipei City, Taiwan; School of Medicine, College of Medicine, Fu Jen Catholic University, New Taipei City, Taiwan; Division of Nephrology, Department of Internal Medicine, Fu Jen Catholic University Hospital, Fu Jen Catholic University, New Taipei City, Taiwan; Koo Foundation Sun Yat-Sen Cancer Center, Taipei, Taiwan; Division of Urology, Taipei City Hospital Ren Ai Branch, Taipei, Taiwan; University of Taipei, General Education Center, Taipei, Taiwan; Department of Urology, College of Medicine and Shu-Tien Urological Research Center, National Yang-Ming Chiao Tung University, Taipei, Taiwan; Division of Gastroenterology and Hepatology, Department of Internal Medicine, Ditmanson Medical Foundation Chia-Yi Christian Hospital, Chiayi City, Taiwan; Clinical Trial Center, Department of Medical Research, Ditmanson Medical Foundation Chia-Yi Christian Hospital, Chiayi City, Taiwan; Fu-Jen Catholic University School of Medicine, Taiwan; Cathay General Hospital Department of Clinical Pathology, Taipei, Taiwan; Cathay General Hospital Department of Internal Medicine, Taipei, Taiwan; School of Medicine, Fu-Jen Catholic University, New Taipei City, Taiwan; Department of Anesthesiology, Cathay General Hospital, Taipei, Taiwan; Department of Urology, Fu Jen Catholic University Hospital, Fu Jen Catholic University, New Taipei City, Taiwan; Department of Medical Research, Far Eastern Memorial Hospital, New Taipei City, Taiwan; Graduate Institute of Medicine and Graduate Program in Biomedical Informatics, Yuan Ze University, Taoyuan, Taiwan; Rheumatology, Immunology and Allergy, Tri-Service General Hospital, Taipei, Taiwan

## Abstract

Incorporating pharmacogenetics into clinical practice promises to improve therapeutic outcome by choosing the medication and dosage optimized for a patient based on genetic factors that affect drug response^1^. One of the most promising benefits of PGx-guided therapy is the avoidance of adverse reactions^2^. To evaluate the clinical impact of PGx risk variants on adverse outcomes, we performed a retrospective study and analyzed the genetic and clinical data from the largest Han Chinese cohort assembled by the Taiwan Precision Medicine Initiative. We found that nearly all participants carried at least one genetic variant that could affect drug response, with many carrying multiple risk variants. Here we show that detailed analyses of four gene-drug pairs, for which sufficient data exist for statistical power, validate previous findings that PGx risk variants are significantly associated with drug-related adverse events or ineffectiveness. However, the excess risk of side effects or lack of efficacy is small compared to that found in those without the PGx risk variants, and most patients with PGx variants do not suffer from adverse events. Our results point to the need for identifying additional risk factors that cause adverse events in patients without PGx risk variants and factors that protect those with PGx risk variants from adverse events.

## INTRODUCTION

Variability in drug effectiveness or safety greatly impacts therapeutic outcome, with drug-response rates varying widely from 25% to 80% among the commonly used medications^3^. Twin and case/control studies have established that genetic factors contribute to variability in drug response. Pharmacogenetics (PGx) explores the influence of an individuals’ genetic makeup on drug metabolism, efficacy, and adverse effects^1^. Incorporating PGx into clinical practice is a promising strategy for healthcare providers to tailor medication selection and dosing to maximize therapeutic benefits while minimizing the risk of drug-related adverse events^1^. PGx also gains increasing attention in pharmaceutical industry for its potential in drug development and drug repositioning^4^.

To facilitate the clinical implementation of PGx, experts from regulatory agencies and consortia such as the US FDA (Food and Drug Administration)^5,6^ and CPIC (Clinical Pharmacogenetics Implementation Consortium)^7^ published clinical practice recommendations based on level of evidence and advocated for incorporating preemptive pharmacogenetic testing into routine care. In a comprehensive study examining PGx variation within the UK Biobank, it was found that the average participant carried genetic variants that would affect their response to ∼10 medications based on CPIC guidelines^7^. In Australia, merely 4% of the study participants lacked actionable PGx variants, and 42% of them had more than 2 actionable PG variants^8^. These findings argue for large-scale implementation of PGx-guided therapy. However, the discordant recommendations from different agencies and consortia make clinical implementation of PGx challenging^9-11^. Furthermore, the benefits of PGx-guided therapy have not been established in large, population-based studies, especially in non-European populations. Therefore, it is crucial to perform studies of large cohorts to evaluate the influences of PGx variants before clinical implementation.

The Taiwan Precision Medicine Initiative (TPMI), a consortium of researchers from the Academia Sinica and 33 partner hospitals across Taiwan, has enrolled 486,956 participants and obtained genetic and longitudinal clinical data from each person (Yang et al., The Taiwan Precision Medicine Initiative: A Cohort for Large-Scale Studies. Submitted to BioRxiv, DOI pending.). With access to their drug prescription and drug-related adverse event history on several commonly prescribed medications, we conducted a retrospective study to analyze four PGx gene-drug pairs with dosing guidelines from CPIC and FDA to determine the impact of PGx risk variants on drug response and toxicity in the largest Asian cohort ever studied in pharmacogenetics. Specifically, we evaluated the association between genetic variants and risk for adverse events (including azathioprine-induced myelosuppression, clopidogrel-related major cardiovascular events (MACEs), statin-associated myopathy (SAMs), and NSAID-linked gastrointestinal (GI) and renal toxicity) based on drug prescription history, clinical intervention, and laboratory test results.

## RESULTS

### Landscape of clinically actionable pharmacogene variants in the cohort

The 486,956 Han Chinese participants of the TPMI were genotyped with one of two SNP arrays (TPMv1 with 686,463 SNPs or TPMv2 with 743,227 SNPs) that contain 3,949 and 2,911 PGx markers, respectively. In this study, we extracted for the cohort the risk variant (star alleles) status of clinically actionable PGx markers or HLA types in 19 pharmacogenes together with the associated phenotypes, including their effects on metabolic enzymes, transporters, immune mediators, and mitochondria proteins (Table s1, s2, and Figure 1a). Variants in these 19 genes affect the response of 58 commonly prescribed medication. Overall, 99.9% of TPMI participants possess at least one PGx variant, which is mainly due to the highly prevalent VKORC1 rs9923231 (–1639G>A) variant, as previously seen in other Han Chinese cohorts ^12,13^. On average, each TPMI participant carries 4.3 clinically actionable PGx risk variants (Figure 1b).

**Figure 1.**
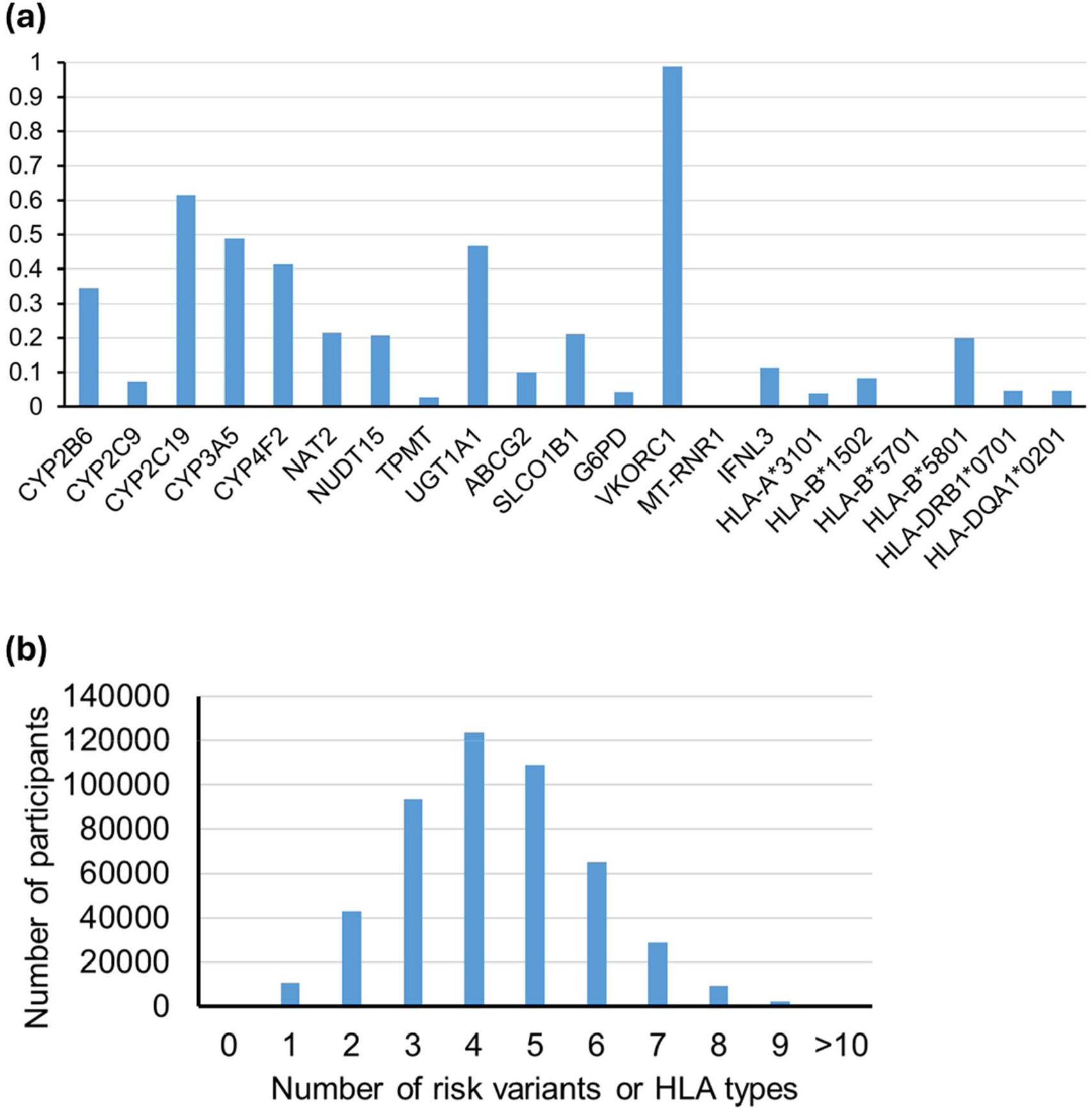
TPMI participants with actionable variants/HLA types in 19 pharmacogenes. (a)The fraction of TPMI participants with actionable variants/HLA types in pharmacogenes; (b)The number of individuals carrying actionable PGx risk variants or HLA types.

### Medication use in people carrying PGx

We extracted and analyzed the drug prescription data from the electronic medical record (EMR) of TPMI participants to determine their medication usage. 48.7% of the TPMI participants have been prescribed at least one of the 58 drugs with prescription guidelines based on their genetic status in the 19 pharmacogenes. 28.4% took two or more drugs with PGx information (Table s3). Individuals with CYP2C19 loss-of-function (LoF) alleles and SLCO1B1 decreased or poor function alleles have been exposed to more drugs with prescription guidelines. Among the individuals carrying clinically actionable PGx variants, 17.8% of them have been prescribed the responding high-risk drugs (Figure 2a and Table s2). The top 20 drugs prescribed include statins, non-steroidal anti-inflammatory drugs (NSAIDs), proton-pump inhibitors (PPI), anti-platelet drugs, and antibiotics (Table 1).

**Table 1.** Clinical practice recommendations for the TPMI participants who took the drug based on their PGx phenotype.

| Drug | Total | Standard treatment | Increased dose | Decreased dose | Alternative | Enzyme activity test | Used in caution | Indetermined |
| --- | --- | --- | --- | --- | --- | --- | --- | --- |
| Abacavir | 692 | 691 |  |  | 1 |  |  |  |
| Abrocitinib | 11 | 9 |  | 2 |  |  |  | - |
| Allopurinol | 5,251 | 4,373 |  |  | 878 |  |  |  |
| Amikacin | 635 | 631 |  |  | 3 |  |  | 1 |
| Atazanavir | 56 | 50 |  |  | 6 |  |  | - |
| Atorvastatin | 70,273 | 55,516 |  | 14,752 |  |  |  | 5 |
| Azathioprine | 8,486 | 6,602 |  | 1,774 | 72 |  |  | 38 |
| Brivaracetam | 28 | 25 |  | 3 |  |  |  | - |
| Carbamazepine | 1,319 | 1,210 |  |  | 109 |  |  |  |
| Celecoxib | 50,200 | 46,570 |  | 3,608 |  |  |  | 22 |
| Citalopram | 6,942 | 2,714 |  | 4,160 | 51 |  |  | 17 |
| Clobazam | 1,283 | 484 |  | 799 |  |  |  | - |
| Clopidogrel | 33,664 | 13,101 |  | 20,517 |  |  |  | 46 |
| Dapsone | 108 | 96 |  |  | 2 | 5 |  | 5 |
| Dexlansoprazole | 19,147 | 7,387 |  | 11,604 | 122 |  |  | 34 |
| Doxepin | 6,773 | 5,745 |  |  | 1,018 |  |  | 10 |
| Efavirenz | 258 | 179 |  | 79 |  |  |  | - |
| Escitalopram | 6,845 | 2,677 |  | 4,100 | 51 |  |  | 17 |
| Flurbiprofen | 28,989 | 26,807 |  | 2,159 |  |  |  | 23 |
| Fluvastatin | 3,526 | 2,548 |  | 968 | 5 |  |  | 5 |
| Gentamicin | 23,953 | 23,816 |  |  | 109 |  |  | 28 |
| Ibuprofen | 6,681 | 6,135 |  | 541 |  |  |  | 5 |
| Imipramine | 9,636 | 8,195 |  |  | 1,420 |  |  | 21 |
| Irinotecan | 596 | 324 |  | 264 |  |  |  | 8 |
| Isoniazid | 2,918 | 2,269 |  |  |  |  | 646 | 3 |
| Kanamycin | 5 | 5 |  |  | - |  |  | - |
| Lansoprazole | 44,378 | 17,063 | 284 | 26,955 |  |  |  | 76 |
| Lapatinib | 27 | 27 |  |  |  |  |  |  |
| Lovastatin | 3,563 | 2,784 |  | 779 |  |  |  | - |
| Meloxicam | 10,174 | 9,447 |  | 715 | 7 |  |  | 5 |
| Mercaptopurine | 37 | 30 |  | 6 | - |  |  | 1 |
| Nateglinide | 227 | 227 |  | - |  |  |  | - |
| Nilotinib | 73 | 66 |  |  |  |  | 6 | 1 |
| Nitrofurantoin | 5 | 4 |  |  | - | 1 |  | - |
| Omeprazole | 33,407 | 12,807 | 210 | 20,344 |  |  |  | 46 |
| Oxcarbazepine | 2,229 | 2,056 |  |  | 173 |  |  |  |
| Pantoprazole | 28,138 | 10,889 | 185 | 17,024 |  |  |  | 40 |
| Paromomycin | 15 | 15 |  |  | - |  |  | - |
| Pazopanib | 120 | 120 |  |  |  |  |  |  |
| Peginterferon alfa-2a | 576 | 504 |  |  |  |  | 72 |  |
| Peginterferon alfa-2b | 218 | 181 |  |  |  |  | 37 |  |
| Phenytoin | 670 | 574 |  | 42 | 54 |  |  | - |
| Piroxicam | 13,933 | 12,841 |  | 1,061 | 17 |  |  | 14 |
| Pitavastatin | 30,876 | 24,284 |  | 6,591 |  |  |  | 1 |
| Pravastatin | 6,955 | 5,510 |  | 1,445 |  |  |  | - |
| Rasburicase | 31 | 29 |  |  | 1 | - |  | 1 |
| Ribavirin | 1,873 | 1,591 |  |  |  |  | 282 |  |
| Rosuvastatin | 46,617 | 17,019 |  | 5,263 |  |  | 24,320 | 15 |
| Sertraline | 4,786 | 3,895 |  | 868 | 12 |  |  | 11 |
| Simvastatin | 10,230 | 7,973 |  | 2,256 |  |  |  | 1 |
| Streptomycin | 59 | 59 |  |  | - |  |  | - |
| Sulfamethoxazole and Trimethoprim | 7,699 | 6,016 |  |  |  |  | 1,666 | 17 |
| Sulfasalazine | 9,777 | 7,679 |  |  |  |  | 2,082 | 16 |
| Tacrolimus | 3,299 | 1,668 | 1,630 |  |  |  |  | 1 |
| Tenoxicam | 23 | 23 |  | - | - |  |  | - |
| Tobramycin | 4,190 | 4,168 |  |  | 16 |  |  | 6 |
| Voriconazole | 343 | 292 |  |  | 51 |  |  |  |
| Warfarin | 6,111 | 1,203 |  | 4,905 |  |  |  | 3 |

**Figure 2.**
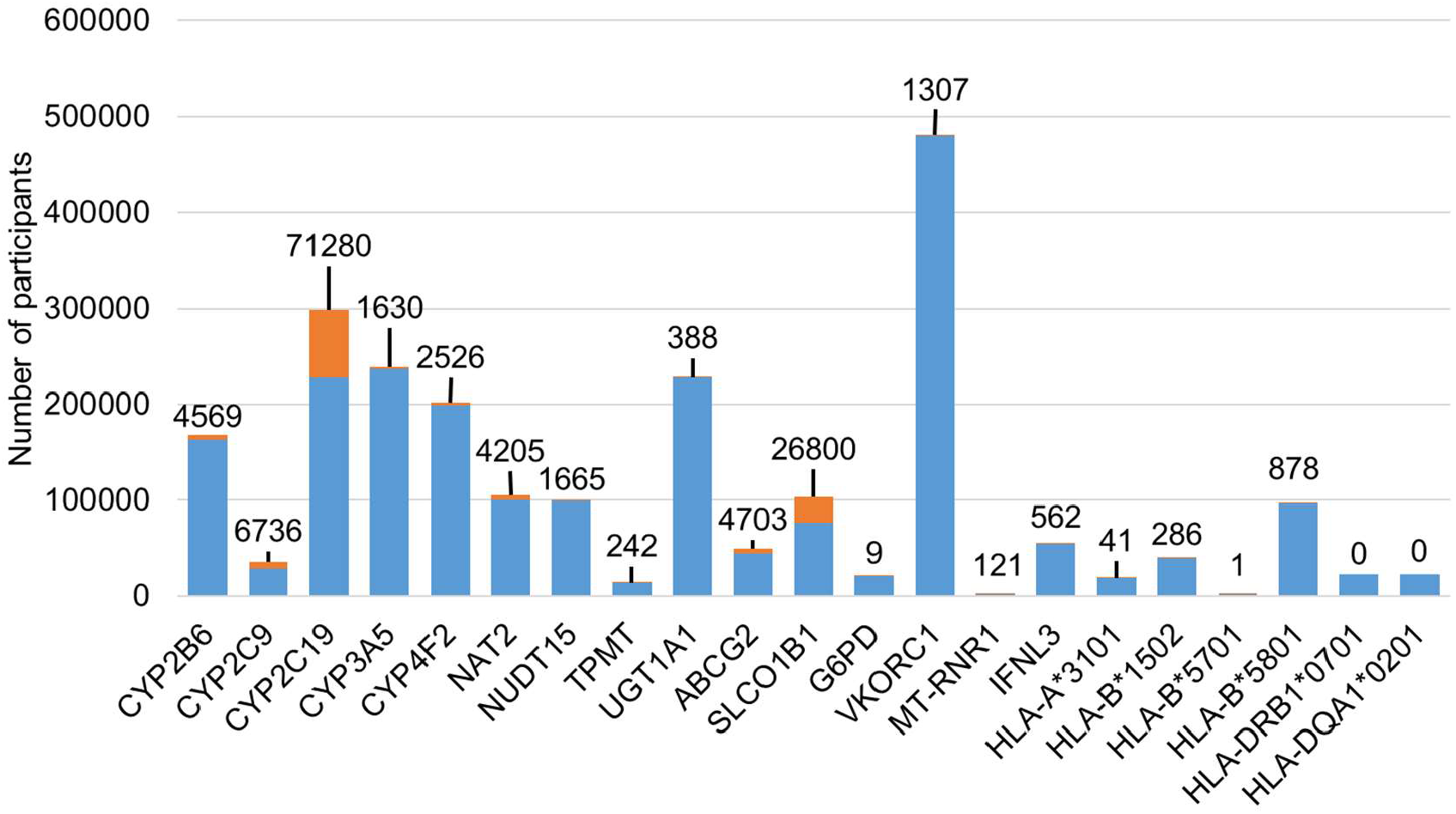
The distribution of risk drugs prescription in people carrying actionable PGx variants. The number of people carrying actionable PGx risk variants or HLA types who took (orange) or did not take (blue) the drug for which they are at risk; number above bar denotes those who took the drug for which they are at risk.

Based on the PGx clinical practice recommendations from US FDA and CPIC, those carrying actionable PGx variants need to have adjusted dosage, to be given alternative drugs, or to be paid extra attention when they are prescribed with high-risk drugs (Table 1). Because the PGx risk variant status of these individuals was unknown when they were prescribed the medication, the PGx guidelines were not followed. However, based on the available medical data, most patients did not suffer from any of the predicted adverse effects.

### Treatment outcomes of those with PGx risk variants/HLA types

We examined the clinical outcomes of TPMI participants who were prescribed drugs known to be affected by PGx variants/HLA types by assessing their EMRs for the drug prescribed, any dosage/drug changes or clinical intervention, and lab test results. By comparing the therapeutic outcomes of patients with or without clinically actionable PGx variants/HLA types, we aim to determine the impact of PGx screening in reducing adverse drug events and improving therapeutic responses. We selected 4 gene-drug pairs for which the TPMI data had large enough sample size and comprehensive clinical data for outcome assessment.

### Impact of CYP2C19 risk variants on clopidogrel-related major adverse cardiovascular events (MACE)

Clopidogrel is an anti-platelet drug commonly used in patients with unstable angina (UA), myocardial infarction (MI), stroke, and peripheral arterial disease to prevent major adverse cardiovascular events (MACE). Clopidogrel is a prodrug that requires enzymatic activation, primarily through CYP2C19. A meta-analysis showed that clopidogrel exhibited a significantly higher risk of MACE in patients with CYP2C19 loss-of-function (LOF) alleles^14^.

Data from 28,055 clopidogrel users who met the criteria for inclusion were used in for this study (Figure s1). Among them, 12.4% developed MACE, including myocardial infarction (MI, 2.1%), unstable angina (UA, 2.5%), heart failure (HF, 4.7%), target lesion revascularization (TLR, 3.7%), stroke (2.3%), or cardiovascular death (CV death, 0.4%). The demographics, clinical characteristics, and CYP2C19 status of the patients are found in Table s4, and there is no significant difference in age, gender, and comorbidities between individuals with or without MACE. Of note, concomitant use of the proton pump inhibitors showed significant association with clopidogrel-related MACE (35.3% vs. 40.8%, P =1.6×10^−10^), which aligns with previous findings from a nationwide population-based study using the Taiwan National Health Insurance database^15^”.

Patients taking clopidogrel with either one or two CYP2C19 LoF alleles were significantly associated with MACE compared to those with non-LoF alleles (P = 2.97 × 10^−27^, OR = 1.53, 95% CI = 1.42–1.65), including MI (P = 2.09 × 10^−31^, OR = 3.98, 95% CI = 3.15–5.02), UA (P = 7.24 × 10^−13^, OR = 1.88, 95% CI = 1.58–2.23), HF (P = 1.77 × 10^−4^, OR = 1.37, 95% CI = 1.22– 1.55), TVR (P = 1.91 × 10^−4^, OR = 1.29, 95% CI = 1.13–1.47), and stroke (P = 6.31 × 10^−5^, OR = 1.41, 95% CI = 1.19–1.67) under multivariate analysis (Figure 3). Both CYP2C19 IM (intermediate metabolizer) and PM (poor metabolizer) had higher MACE incidence under clopidogrel treatment; however, 85.9% of people with CYP2C19 LOF alleles tolerated clopidogrel treatment well. There is no significant difference in risk of CV death under clopidogrel therapy for those with or without CYP2C19 LoF alleles (Figure 3).

**Figure 3.**
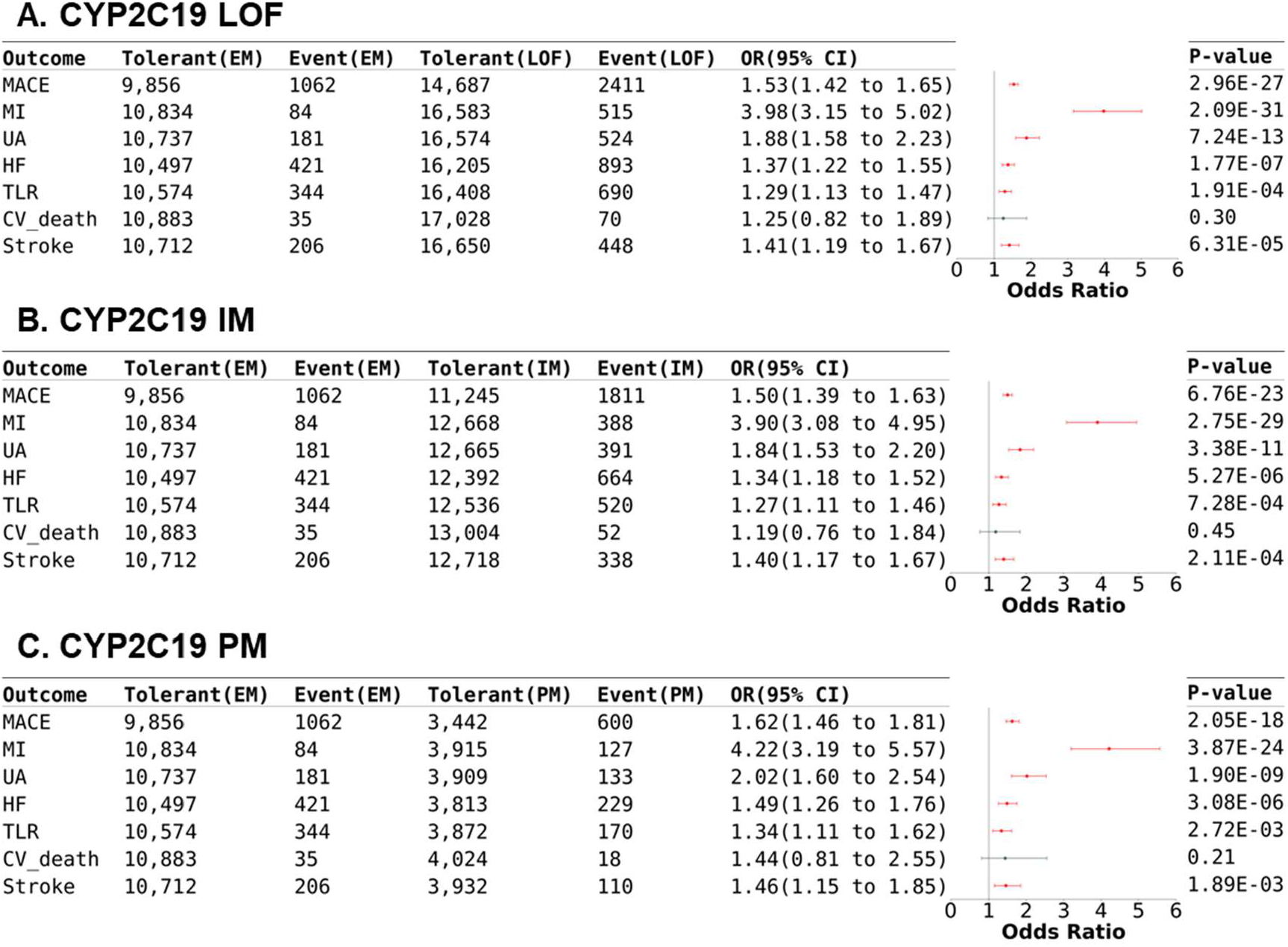
Impact of CYP2C19 in clopidogrel-related MACE. Forest plot of the MACE risk in clopidogrel users with different CYP2C19 phenotypes. The significant associations are in red. CV_death = cardiovascular death; HF = heart failure; IM = intermediate metabolizer; LoF = loss-of-function; MACE = major adverse cardiovascular events; MI = myocardial infarction; PM = poor metabolizer; TLR = target lesion revascularization; UA = unstable angina.

### Impact of NUDT15/TPMT in azathioprine-related adverse events

Azathioprine (AZA) is a commonly prescribed immunosuppressive antimetabolite in the management of acute lymphoblastic leukemia, autoimmune conditions, and organ transplantation. AZA has a narrow therapeutic index and a high potential for adverse drug reactions, like bone marrow toxicity, hepatotoxicity, massive hair loss, nausea and vomiting. Nudix hydrolase 15 (NUDT15) and thiopurine-S-methyltransferase (TPMT) are two important enzymes that decrease the concentration of AZA active metabolites, so higher AZA toxicity risk is commonly observed in people with deficient NUDT15 and TPMT^16-18^. Both CPIC and US FDA recommend a significant or substantial dose reduction for NUDT15 or TPMT intermediate or poor metabolizers (IM and PM)^6,7^.

We studied 8,451 participants using azathioprine for prevention of renal transplant rejection and treatment of inflammatory conditions, including systemic lupus erythematosus (SLE), Sjogren syndrome, rheumatoid arthritis (RA), other connective tissue disorders, Crohn’s disease, ulcerative, or severe atopic dermatitis (Figure s2). A total of 1,503 (17.8%) patients stopped azathioprine treatment due to intolerable adverse drug reactions such as leukopenia (10.7%), thrombocytopenia (6.8%), hepatitis (4.6%), GI discomfort (1%), alopecia (0.7%), and allergy (0.6%). The distribution of sex, comorbidity, and most of concurrent medication usage were not significantly different in the patients with AZA-induced adverse reactions (ADRs) compared with AZA tolerant controls, but concomitant use of allopurinol and AZA increased risk for adverse events development (Table s5). The age of initial use of AZA in the ADR group is slight younger than the tolerant control group.

The NUDT15 IM (P = 5.44 × 10^−4^, OR = 1.28, 95% CI = 1.11–1.48) and NUDT15 PM (P = 2.88 × 10^−8^, OR = 4.05, 95% CI = 2.47–6.64) phenotypes were associated with AZA-induced adverse events under multivariant regression analysis (Figure 4A-C). The incidences of leukopenia in intermediate/poor NUDT15 metabolizers (14.9% and 37.7%, respectively) patients were significantly higher than that in extensive NUDT15 metabolizers (EM; 9.4%) (P = 4.47×10^−14^, OR = 1.85, 95% CI = 1.58–2.17). The significant associations between azathioprine discontinuation due to thrombocytopenia (P = 4.91×10^−3^, OR = 2.67, 95% CI = 1.35–5.3) or massive hair loss (P = 1.24×10^−5^, OR = 10.84, 95% CI = 3.72–31.58) and NUDT15 were only observed in poor metabolizers. AZA discontinuation rates due to adverse events did not differ between patients with normal and decreased TPMT activity (Figure 4 D).

**Figure 4.**
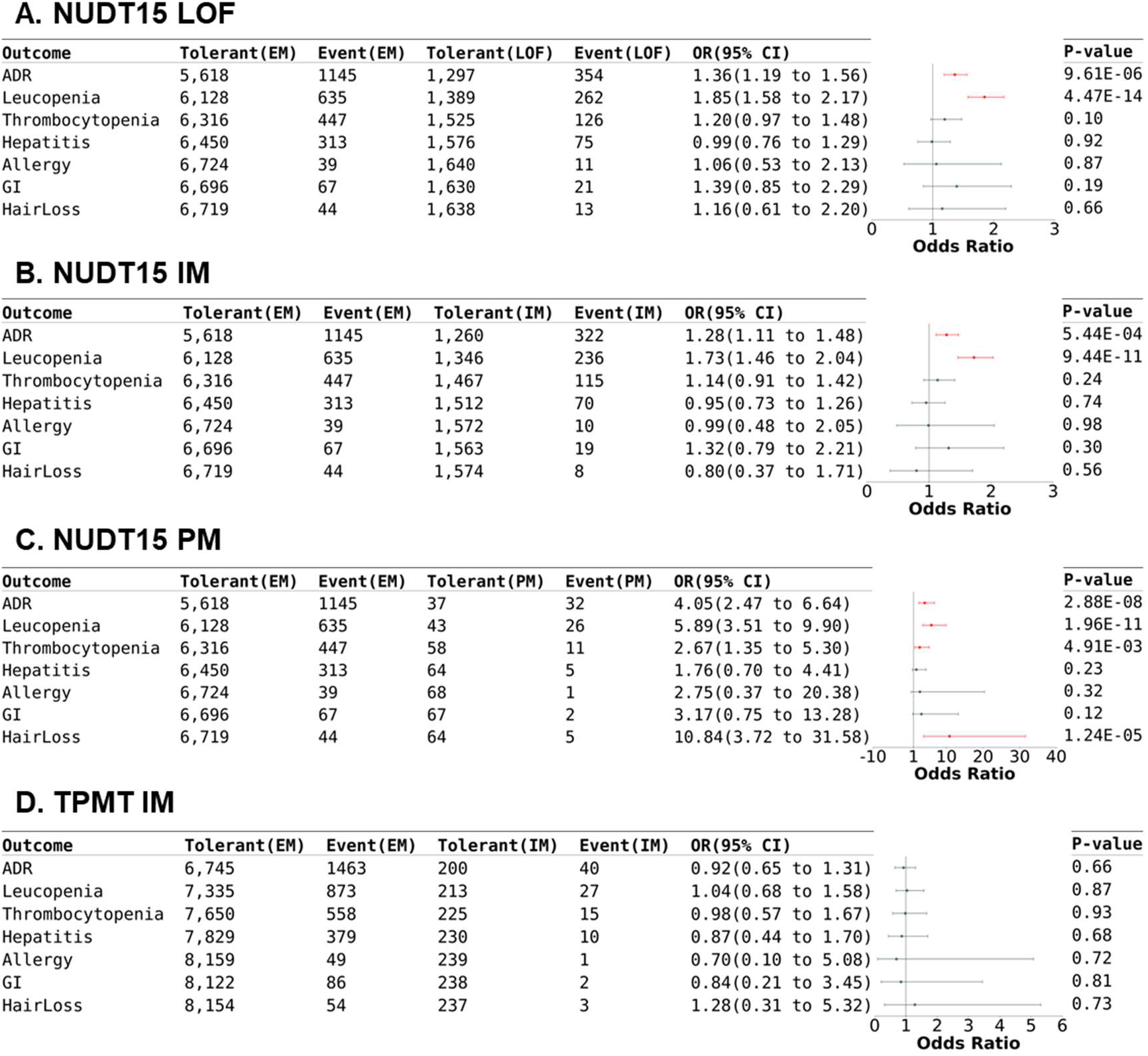
Influence of NUDT15 and TPMT for azathioprine discontinuation due to ADR. Forest plot of the ADR risk in azathioprine users with different NUDT15 and TPMT phenotypes. The significant associations are in red. ADR = adverse reactions; GI = gastrointestinal discomfort; IM = intermediate metabolizer; LoF = loss-of-function; PM = poor metabolizer.

Again, the clinical impact is limited, with only 21.4% of NUDT15 IM/PM and 16.7% of TPMT IM had severe AZA-induced ADRs, whereas 16.9% of NUDT15 EM and 17.8% of TPMT EM suffered from AZA-induced ADRs.

### Impact of ABCG2/CYP2C9/SLCO1B1 in statin-associated myopathy

Statins are used in reducing plasma low-density lipoprotein cholesterol (LDL-C) levels and preventing the risk of atherosclerotic cardiovascular diseases^19^. However, statins use is often associated with side-effects, particularly those related to musculoskeletal and hepatic systems. Statin-associated muscle events (SAMs) stand out as the most frequently reported side effects, encompassing a spectrum of clinical presentations ranging from mild symptoms, such as muscle pain and weakness, to severe muscle injury, such as rhabdomyolysis^20^. Establishing the causal relationship between muscle complaints and statin use is challenging, particularly in the case of subjective symptoms such as myalgia^21,22^. To mitigate the risk of SAMs over-diagnosis, we reviewed and extracted EMR data on symptom relief following statin withdrawal or symptom recurrence upon statin rechallenge. Data from 127,197 TPMI participants who ever treated with any type of statins (atorvastatin, fluvastatin, lovastatin, pitavastatin, pravastatin, rosuvastatin, or simvastatin) were analyzed for this study (with individuals previously suffering from muscular disorders excluded). 34,411 participants took more than one statin, and 7 were treated with 6 types of statins (Table s6). Although statins shared similar structure, many participants who suffer from muscle toxicity caused by one statin can take another statin without any side effects (Table s6). Therefore, genetic associations with myopathy induced by different statins were analyzed separately (Figure 5 and Table s7-s9).

**Figure 5.**
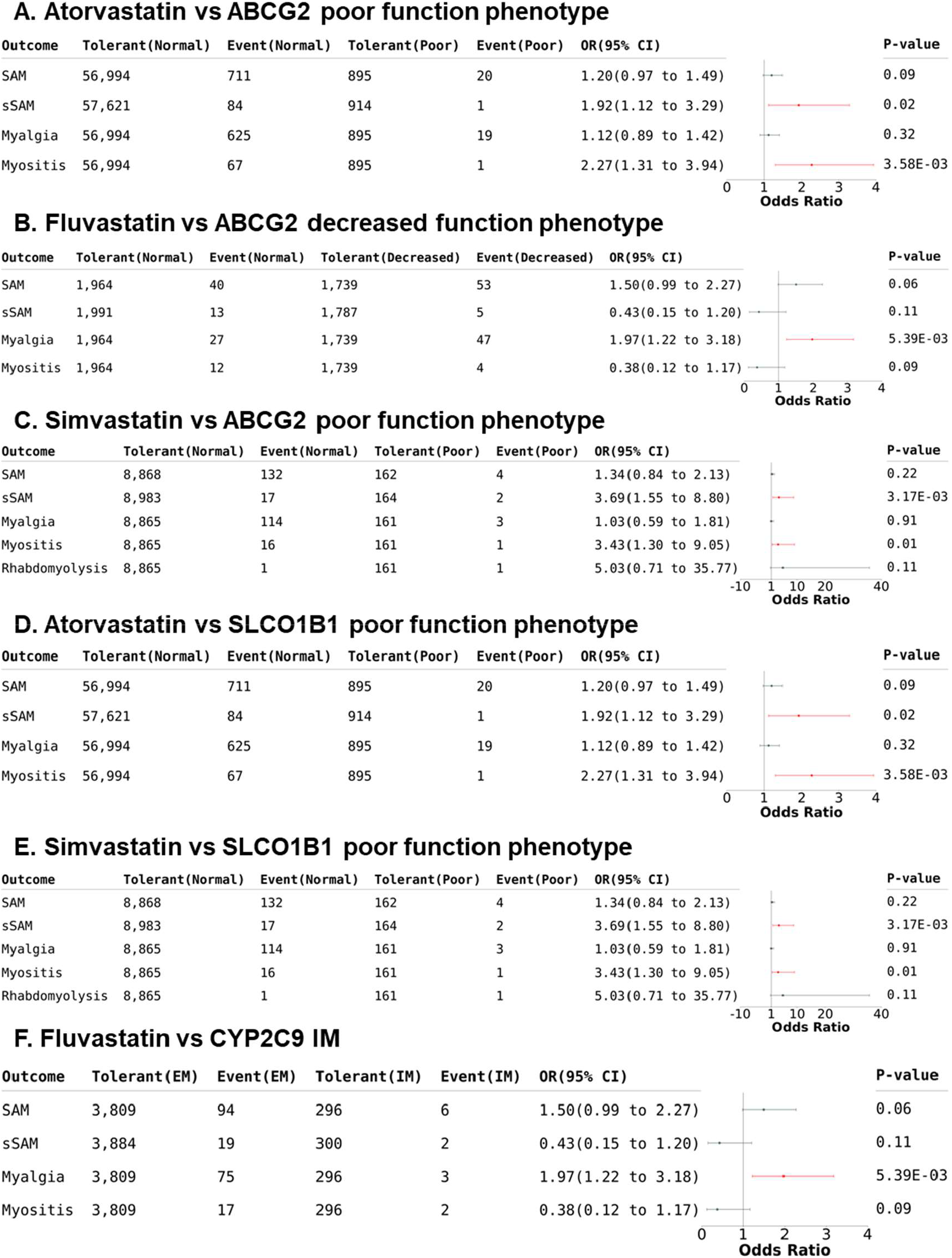
Influence of ABCG2/SLCO1B1/CYP2C9 in statin-associated myopathy. Forest plot of the SAM risk in statin users with different ABCG2, SLCO1B1, or CYP2C9 phenotypes. The significant associations are in red. SAM = statin-associated muscle events; sSAM = severe forms of statin-associated muscle events.

Our study shows that people with poor function of ABCG2 have higher risk of myositis when taking atorvastatin (P = 3.58×10^−3^, OR = 2.27, 95% CI = 1.31–3.94) or simvastatin (P = 1.1×10^−2^, OR = 3.43, 95% CI = 1.3–9.05); whereas people with ABCG2 decreased or poor function phenotype have higher risk of myalgia while taking fluvastatin (P = 5.39×10^−3^, OR = 1.97, 95% CI = 1.22–3.18) (Figure 5A-C and Table s7). Patients with SLCO1B1 poor function phenotype (c.521T>C) have increased risk in developing severe myositis with atorvastatin (P = 3.58×10^−3^, OR = 2.27, 95% CI = 1.31–3.94) and simvastatin (P = 1.26×10^−2^, OR = 3.43, 95% CI = 1.3–90.5) (Figure 5D-E and Table s8). Furthermore, the frequency of CYP2C9 IM is significantly higher in patients who experience myalgia after receiving fluvastatin (P = 5.39×10^−3^, OR = 1.97, 95% CI = 1.22–3.18) (Figure 5F, Table s9).

Despite the significant association in some of the ADEs, the frequencies are so low that the vast majority of the “high risk” individuals (98% or more) do not suffer from any ADEs.

### Impact of CYP2C9 in NSAID-associated adverse events

Nonsteroidal anti-inflammatory drugs (NSAIDs) are known for their ability to reduce pain, inflammation, and fever by inhibiting the production of prostaglandins. Although substantial evidence links CYP2C9 deficient phenotype to plasma NSAID concentrations, an increased risk of ADEs in individuals with reduced CYP2C9 metabolism of NSAIDs (such as celecoxib, flurbiprofen, ibuprofen, lornoxicam, meloxicam, piroxicam and tenoxicam) has not be substantiated. However, since the NSAID toxicity is dose- and duration-dependent^23^, a recommendation for NSAIDs dose adjustment and selection based on CYP2C9 genotype was issued by CPIC^24^. To assess the impact of CYP2C9 in NSAID-related ADEs, we analyzed the TPMI data to evaluate the association between CYP2C9 activity and NSAID-related upper gastrointestinal (GI) and renal events. We find that those with the CYP2C9 PM phenotype is predisposed to NSAID-induced upper GI bleeding, although the sample size is relatively small (Table s11). It is important to note that comorbidities and concurrent medications contribute much more to the side effects of NSAIDs than CYP2C9 status (Table s10).

## DISCUSSION

The benefits of pharmacogenetic testing in preventing severe adverse reactions are well documented^1^. Several randomized controlled trials (RCTs) and studies have highlighted the benefit of PGx-guided therapy, showcasing its potential in optimized medication selection and dosing that leads to improved efficacy and safety^25-28^. However, these small, mostly European studies do not address the applicability of PGx-guided therapy in non-European populations such as the Han Chinese, where literally everyone has PGx risk variants that affect his/her response to drugs developed with clinical trials conducted mostly with subjects of European ancestry. Differences in drug response across populations are well-recognized, leading to distinct optimal doses between Asians and Europeans in current clinical practice. Therefore, the dose limitation listed in PGx dosing guideline should be optimized based on ethnicity. For instance, the US FDA notes a higher risk of myopathy in patients with decreased or poor function of SLCO1B1 when taking 80mg of simvastatin^6^. CPIC recommends a daily dosage of simvastatin below 20mg for patients with decreased function of SLCO1B1^7^. However, in Taiwan, the most prescribed and effective dose of simvastatin ranges from 10-20mg, regardless of SLCO1B1 genotype^29^.^30,31^

Our retrospective study of the 4 gene-drug pairs in 486,956 participants of the TPMI, for whom we have both genetic and longitudinal clinical data, shows that PGx-guided therapy is not straightforward. Firstly, while our findings validate the published results that PGx risk variants increase the risk of adverse events, and the increase is statistically significant, the relative risk is low or moderate. Secondly, the vast majority of those with PGx risk variants who take the drug in question do not suffer from the predicted side-effects. Conversely, a significant fraction of those without PGx risk variants suffer from adverse reactions. Thirdly, many of the adverse reactions are reversible, non-life-threatening events, and can be managed easily. Finally, as there are no equally effective drug alternatives available in many cases, so to avoid adverse reactions by not taking a medication means that one is taking a medication of lower efficacy.

Given our findings, one must proceed with caution when implementing PGx-guided therapy. Further studies must be conducted to (1) identify additional (genetic and non-genetic) factors that cause adverse drug reactions that explain the baseline occurrence of such events in those without PGx risk variants; (2) identify factors that protect those with PGx risk variants from adverse effects while taking the medication in question; and (3) evaluate risk-management strategies for PGx risk variant carriers that allow them to use the most effective therapy even if they are at risk for adverse reactions by closely monitoring the emergence of side effects.

Although our findings and conclusions are strong for the 4 gene-drug pairs studied, and the results can be extrapolated to other medications, our study has several limitations. First, the TPMI obtains clinical data for each participant from the hospital through which he/she is enrolled, clinical data from other healthcare providers for the participant are not part of the dataset, leading to the possibility of under-reporting. Second, the genetic data are based on SNP-array data, which means that not all the PGx risk variants in each pharmacogene are represented. For example, some variants are embedded in repeat regions that no suitable probes can be designed on the array. In addition, variants of extremely low allele frequency (<0.1% minor allele frequency) cannot be typed accurately on the array and are therefore excluded from the design. Third, the clinical phenotypes are extracted from the clinical data based on chart-review and the variability in clinical note style and substance across hundreds of doctors from 33 hospitals create noise in the data. Some cases and controls are likely excluded due to this reason. Apart from genetic variants, the heterogeneity in drug response may also stem from other factors, including environmental and nutritional influences, disease severity, comorbidities, concomitant medications, and individual patient lifestyles. To overcome these limitations, future studies will be conducted with comprehensive clinical data from the national health insurance database (that collects information from all hospitals and clinics the participants obtain their care), comprehensive genetic data from whole genome sequencing, and prospective study with standardized recording of clinical outcomes. Finally, the current study does not include analysis of the risk variants in CYP2D6, the gene that encodes a pivotal enzyme in the metabolic pathways of nearly 20% of frequently prescribed medications^32^. The challenge of accurately typing CYP2D6 is due to the high degree of polymorphism and complex nature with structural variations (SV) in the gene^33^. The lack of multiplex PCR and SV analysis algorithm make accurately determining CYP2D6 status from SNP array data impossible at this time. An optimal pipeline to overcome this challenge is under construction.

In conclusion, we report results from the largest retrospective study in non-Europeans on the distribution of PG risk variants and their impact for 4 widely accepted gene-drug pairs on drug-related adverse reactions and treatment responses. Our findings show that implementing PGx-guided therapy in large populations is not simple and should be done with caution. We speculate that these conclusions apply to many other gene-drug pairs and to non-European populations, highlighting the need for comprehensive, integrative strategies to enhance the safety and efficacy of precision medicine.

## METHODS

### Data source

This study was conducted with genetic and clinical datasets from the Taiwan Precision Medicine Initiative (TPMI) cohort, with participants recruited from 16 medical centers (encompassing 33 hospitals) in Taiwan. Genetic data include the whole genome genotyping data on custom-designed TPMI SNP arrays, TPM and TPM2 array, genome-wide imputation data, and human leukocyte antigen (HLA) imputation data. The EMRs were provided by the hospital through which the participants were enrolled. They included outpatient, inpatient, and emergency room (ER) visiting records, medication prescription records, discharge summaries and operation notes, laboratory test results, and reports of pathology, surgery, image, and Mini-Mental State Examination (MMSE). The 1,498 whole genome sequencing (WGS) data from the Taiwan Biobank were used for the imputation reference panel and for genotype validation^34^. The DeepVariant variant calling pipelines were used, and SHAPEIT5 and IMPUTE5 were applied for phasing and imputation^35-37^ (Yang et al., The Taiwan Precision Medicine Initiative: A Cohort for Large-Scale Studies. Submitted to BioRxiv, DOI pending.). For the imputation of G6PD, which is located on chromosome X, we utilized a reference panel derived from samples of TPM2 due to the absence of relevant data points in TPM1. The imputation process was conducted using Shapeit4 and Impute5. Male samples were imputed with the --haploid parameter in Impute5 to account for the haploid nature of the X chromosome in males, while female samples underwent standard diploid imputation. The training and validating dataset for Hibag encompassed 1,359 HLA typing data sourced from multiple repositories, including the Adverse Drug Reaction Project, the Collaborative Study to Establish a Cell Bank and a Genetic Database on Non-Aboriginal Taiwanese, and the Taiwan Biobank. Utilizing the Hibag algorithm, classical HLA alleles such as HLA-A, HLA-B, HLA-C, HLA-DRB1, HLA-DQB1, and HLA-DPB1 were imputed. Each HLA allele underwent training with a 500-kb flanking region and utilized 500 classifiers to construct models, ensuring comprehensive coverage and robust predictive accuracy^34^. This study was approved by TPMI’s committees, and the data usage followed the approval from the ethical committees of the Academia Sinica (AS-IRB01-18079), the Taiwan Biobank (TWBR10806-04), and all participating hospitals: Taipei Veterans General Hospital (2020-08-014A), National Taiwan University Hospital (201912110RINC), Tri-Service General Hospital (2-108-05-038),Chang Gung Memorial Hospital (201901731A3), Taipei Medical University Healthcare System (N202001037), Chung Shan Medical University Hospital (CS19035), Taichung Veterans General Hospital (SF19153A), Changhua Christian Hospital (190713), Kaohsiung Medical University Chung-Ho Memorial Hospital (KMUHIRB-SV(II)-20190059), Hualien Tzu Chi Hospital (IRB108-123-A), Far Eastern Memorial Hospital (110073-F), Ditmanson Medical Foundation Chia-Yi Christian Hospital (IRB2021128), Taipei City Hospital (TCHIRB-10912016), Koo Foundation Sun Yat-Sen Cancer Center (20190823A), Cathay General Hospital (CGH-P110041), and Fu Jen Catholic University Hospital (FJUH109001).

### Pharmacogenomic variants analysis

The drug-gene pairs were adopted from Table of Pharmacogenetic Associations, US FDA (https://www.fda.gov/medical-devices/precision-medicine/table-pharmacogenetic-associations) and the Clinical Guideline Annotations table of CPIC guidelines from PharmGkB (https://www.pharmgkb.org/guidelineAnnotations). The drugs with fewer than 10 prescription records were removed from the analysis. The curation of PGX variants and actionable PGx phenotype were based on the instructions from PharmGkB and PharmVar (https://www.pharmvar.org/). The PGx variants were first screened with NGS data to remove the ones with allele frequency lower than 0.1%. Then the SNP genotype and imputation data of passed variants were validated by NGS, Sanger sequencing, and Sequenom MassARRAY platform. The variants with sensitivity and specificity higher than 99% were saved for further analysis. The final list of drug-gene pair evaluated in this study is found in supplemental table S2, and the variants information in supplemental table S1.

### Drug prescription and treatment outcome assessment

When analyzing the number of individual encountered drugs with PGx warnings, 58 approved drugs with clear genetic-base clinical utility guidelines were selected based on their availability in the TPMI dataset. We only kept the individual with two or more prescription records for each drug.

Data from people taking azathioprine (AZA), clopidogrel, statins, and non-steroidal anti-inflammatory drugs (NSAID) metabolized by CYP2C9 were analyzed to assess the influence of PGx variants and phenotypes on drug response.

To study the influence of CYP2C19 on clopidogrel response, clopidogrel users who were 18 years of age or older at initial dose, in good compliance, and without severe allergic events were selected. The endpoint was major adverse cardiovascular event (MACE), including CV death, non-fatal heart failure (HF), non-fatal unstable angina (UA), acute myocardial infarction (MI), acute ischemic stroke or transient ischemic attack (TIA), or target lesion revascularization (TLR) requiring clinical interventions such as percutaneous intervention (PCI) or surgical bypass (extracted from image and operation reports).

The AZA study cohort comprised of AZA tolerant controls and patients who discontinued AZA due to massive hair loss, gastrointestinal discomfort (nausea, vomiting, and diarrhea), allergic reactions, hepatitis (defined as ALS/AST > 3x ULN), leucopenia (defined as WBC < 3,500/mm^3^), and thrombocytopenia (defined as platelet count < 150,000 /uL), as documented by the responsible physicians and/or laboratory test results. We excluded patients with hematological malignancies and poor liver function prior to AZA treatment. The study design is shown in supplementary figure s2.

The TPMI participants taking atorvastatin, fluvastatin, lovastatin, pitavastatin, pravastatin, rosuvastatin, or simvastatin without any muscular disorder history were included to assess the role of ABCG2, CYP2C9, and SLCO1B1 in the risk of statin-associated myopathy (CPK elevation with muscle complaints, myalgia, myositis, and rhabdomyolysis).

Individuals were selected to evaluated the occurrence of adverse renal events and upper GI discomfort in different CYP2C9 phenotype groups if they had prescriptions of NSAIDs mainly metabolized via CYP2C9 (Celecoxib, flurbiprofen, ibuprofen, meloxicam, piroxicam, and tenoxicam) but without history of end stage kidney disease (ESRD, 99.2, N18.5, and N19), poor renal function (<15 ml/min/1.73 m^2^), receiving dialysis (58001C, 58002C, 58009B, 58010B, 58011C, 58012B, 58013C, 58017C, 58018C, and 58026C) a renal replacement therapy (76020B, N26028, T86.1 and Z94.0), alcoholism (F10), esophageal varices (I85 and I98.2), Mallory–Weiss syndrome (K22.6), liver cirrhosis (K70, K72–74, and K76), GI tract cancer (C15, C16, and C17), and coagulation defects (D65-D68).

Comorbidities included diabetes mellitus (E08-E13), hypercholesterolemia (E78.0-E78.5), hypertension (I10), ischemic heart diseases (I20-I25), heart failure (I50), and cerebrovascular diseases (I60-I69) were analyzed as well to identify their influence on treatment outcome.

### Statistical analysis

The frequencies of PGx variants from genotype and NGS data were calculated by PLINK and VCFtools. Fisher exact test and chi-square tests were conducted to compare differences between tolerant and non-tolerant individuals who carry the different PGx variants. Continuous variables were shown as means along with their standard deviations, whereas categorical variables were displayed as numerical counts and percentages. For quantitative data, comparison between 2 groups was performed by 2-tailed student t-test. Logistic regression analysis was utilized to determine the odds ratio (OR) and 95% confidence interval of the allele model. The multivariate logistic regression was adjusted for clinical factors, including sex, age, comorbidities, and concurrent medications. A p value ≤ 0.05 was considered significant in this study.

## Data Availability

The genotyping and electronic medical record data analyzed in this study are from the Taiwan Precision Medicine Initiative with proper approval from the TPMI Data Access Committee. In compliance with the confidentiality laws governing genetic and health data in Taiwan, the de-identified TPMI data are kept in a secure server at the Academia Sinica and not released to the public. Researchers requesting access to the individual genotyping and EMR data can do so on a collaborative basis. Instructions on requesting access to the data can be found on the TPMI official website (https://tpmi.ibms.sinica.edu.tw). In addition to the TPMI data, we analyzed the TWB dataset as part of the validation, and the genotype data from the Taiwan Biobank are available through a formal application process (https://www.twbiobank.org.tw/index.php).

https://www.twbiobank.org.tw/index.php

## DATA AVAILABILITY

The genotyping and electronic medical record (EMR) data analyzed in this study are from the Taiwan Precision Medicine Initiative (TPMI) with proper approval from the TPMI Data Access Committee. In compliance with the confidentiality laws governing genetic and health data in Taiwan, the de-identified TPMI data are kept in a secure server at the Academia Sinica and not released to the public. Researchers requesting access to the individual genotyping and EMR data can do so on a collaborative basis. Instructions on requesting access to the data can be found on the TPMI’s official website (https://tpmi.ibms.sinica.edu.tw). In addition to the TPMI data, we analyzed the TWB dataset as part of the validation, and the genotype data from the Taiwan Biobank are available through a formal application process (https://www.twbiobank.org.tw/index.php).

## COMPETING INTERESTS

The authors declare no competing interests.

## ACKNOWLEDGEMENTS

We thank all the participants and researchers of the Taiwan Precision Medicine Initiative and the Taiwan Biobank. This study was funded in part by the Academia Sinica (40-05-GMM, AS-GC-110-MD02, and 236e-1100202 to P.-Y.K. and J.-Y.W.) and the National Development Fund, Executive Yuan (NSTC 111-3114-Y-001-001 to P.-Y.K.).

## AUTHOR CONTRIBUTION

C.Y.W., P.Y.K., M.S.W., C.K.C., Y.J.S., and J.Y.W. were involved in the conceptualization and design of the study. M.S.L., S.P.W., K.T.L., H.P.C., Y.J.C., J.S., Y.T.C., C.C.C., C.F.K., J.C.L., H.C.K., T.M.C., C.W.L., J.H.L., S.F.L., H.T.C., L.Y.L., L.C.C., C.T.T., H.L.K., J.J.Y., J.S.J., M.C.C., T.C.H., S.F.Y., H.J.L., S.C.S., P.C., P.F.L., C.L.T., C.K.T., S.E.T., C.M.L., Y.F.W., C.Y.H., S.Z.L., C.C.C., T.K.L., S.M.H., C.H.C., C.D.C., G.C.M., T.Y.C., J.J.H., C.L.L., K.J.K., C.F.H., S.S.C., P.Y.C., K.T., C.H.C., C.C.C., H.S.C., Y.L.C., and H.C.C. conducted the investigation. M.F.T., E.C.Y., K.M.C., S.M.C., M.S.L., S.P.W., K.T.L., and Y.T.C. curated the data. C.Y.W., M.F.T., E.C.Y., K.M.C., and S.M.C. performed the formal analysis. T.C.Y., S.L.L., J.Y.W., L.H.L., C.H.C., C.S.-J.F., H.C.Y., Y.T.H., Y.M.L., C.H.C., C.C.C., H.S.C., Y.L.C., H.C.C., and P.Y.K. supervised the project. C.Y.W., and P.Y.K. wrote the initial draft, C.Y.W., J.Y.W., M.F.T., L.H.L., C.H.C., C.S.-J.F., H.C.Y., Y.T.H., H.H.C., Y.M.L., E.C.Y., C.H.C., and P.Y.K. reviewed and edited the manuscript, and all authors were involved in the revision of the manuscript for publication. J.Y.W., C.C.C., H.S.C., Y.L.C., and H.C.C. were involved in the resources. Y.M.L., H.P.C., Y.J.C., J.S., and P.Y.K. were involved in project administration. J.Y.W., Y.M.L., and P.Y.K. were involved in funding acquisition.

## Notes

### Competing Interest Statement

The authors have declared no competing interest.

### Author Declarations

This study was approved by the Institutional Review Boards of Taipei Veterans General Hospital (2020-08-014A), National Taiwan University Hospital (201912110RINC), Tri-Service General Hospital (2-108-05-038), Chang Gung Memorial Hospital (201901731A3), Taipei Medical University Healthcare System (N202001037), Chung Shan Medical University Hospital (CS19035), Taichung Veterans General Hospital (SF19153A), Changhua Christian Hospital (190713), Kaohsiung Medical University Chung-Ho Memorial Hospital (KMUHIRBSV(II)-20190059), Hualien Tzu Chi Hospital (IRB108-123-A), Far Eastern Memorial Hospital (110073-F), Ditmanson Medical Foundation Chia-Yi Christian Hospital (IRB2021128), Taipei City Hospital (TCHIRB-10912016), Koo Foundation Sun Yat-Sen Cancer Center (20190823A) Cathay General Hospital (CGH-P110041), Fu Jen Catholic University Hospital (FJUH109001) and Academia Sinica (AS-IRB01-18079), Taiwan. Written informed consent was obtained from the subjects in accordance with institutional requirements and the Declaration of Helsinki principles. All collected information was de-identified before statistical data analysis.

