## supplementary table and figure for "Clinical Impact of Pharmacogenetic Risk Variants in a Large Chinese Cohort"

**Table s1: List of clinically relevant PGx variants**

| Gene | Allele | Array Type | Data source | Allele Functional Status^ | Activity Score* | TPMI_AF | Sensitivity$ | Specificity$ |
|---|---|---|---|---|---|---|---|---|
| ABCG2 | c.421 C>A | TPM/TPM2 | Genotype | Decreased function | N/A | 31.7% | 99.9% | 100.0% |
| CYP2B6 | decrease variants | TPM/TPM2 | Genotype | Decreased function | N/A | 19.1% | 99.8% | 100.0% |
| CYP2C9 | *3 | TPM/TPM2 | Genotype | Decreased function | 0.5 | 3.2% | 100.0% | 100.0% |
| | *16 | TPM/TPM2 | Genotype | Decreased function | 0.5 | 0.5% | 100.0% | 100.0% |
| CYP2C19 | *2 | TPM/TPM2 | Genotype | No function | N/A | 32.3% | 100.0% | 100.0% |
| | *3 | TPM/TPM2 | Genotype | No function | N/A | 5.2% | 100.0% | 100.0% |
| | *6 | TPM/TPM2 | Genotype | No function | N/A | 0.2% | 100.0% | 100.0% |
| | *17 | TPM/TPM2 | Imputation | Increased function | N/A | 0.5% | 100.0% | 100.0% |
| CYP3A5 | *1 | TPM/TPM2 | Genotype | Normal function | | 28.6% | 100.0% | 100.0% |
| CYP4F2 | *3 | TPM/TPM2 | Genotype | Decreased function | | 23.6% | 100.0% | 100.0% |
| G6PD | Canton/Taiwan-Hakka | TPM/TPM2 | Imputation/Genotype | Deficient function | II | 1.41% | 100.0% | 100.0% |
| | Chinese-3/taipei | TPM/TPM2 | Genotype | Deficient function | III | 0.20% | 100.0% | 100.0% |
| | Chinese-5 | TPM/TPM2 | Genotype | Deficient function | II | 0.25% | 100.0% | 100.0% |
| | Gaohe | TPM/TPM2 | Genotype | Deficient function | II | 0.17% | 100.0% | 100.0% |
| | Kaiping | TPM/TPM2 | Genotype | Deficient function | II | 0.61% | 100.0% | 100.0% |
| | QuingYan | TPM/TPM2 | Genotype | Deficient function | III | 0.17% | 100.0% | 100.0% |
| HLA-A | *31:01 | TPM/TPM2 | HiBag | Risk | | 2.3% | 100.0% | 100.0% |
| HLA-B | *15:02 | TPM/TPM2 | HiBag | Risk | | 4.2% | 100.0% | 99.9% |
| | *57:01 | TPM/TPM2 | HiBag | Risk | | 0.2% | 100.0% | 100.0% |
| | *58:01 | TPM/TPM2 | HiBag | Risk | | 10.6% | 100.0% | 100.0% |
| HLA-DQA1 | *02:01 | TPM/TPM2 | HiBag | Risk | | 2.4% | 100.0% | 100.0% |
| HLA-DRB1 | *07:01 | TPM/TPM2 | HiBag | Risk | | 2.3% | 96.3% | 100.0% |
| IFNL3 | rs12979860 | TPM/TPM2 | Genotype | Unfavorable response | | 5.8% | 99.4% | 100.0% |
| MTRNR1 | m.1095T>C | TPM/TPM2 | Genotype | Risk | | 0.2% | 100.0% | 100.0% |
| | m.1555A>G | TPM/TPM2 | Genotype | Risk | | 0.2% | 100.0% | 100.0% |
| NAT2 | *5 | TPM/TPM2 | Genotype | Decreased Function | | 3.9% | 100.0% | 100.0% |
| | *6 | TPM/TPM2 | Genotype | Decreased Function | | 26.5% | 100.0% | 100.0% |
| | *7 | TPM/TPM2 | Genotype | Decreased Function | | 16.1% | 100.0% | 100.0% |
| NUDT15 | *2/*3 | TPM/TPM2 | Imputation/Genotype | No Function | | 11.0% | 99.7% | 100.0% |
| SLCO1B1 | *5/*15 | TPM/TPM2 | Genotype | No function | | 11.2% | 100.0% | 100.0% |
| TPMT | *3C | TPM/TPM2 | Genotype | No Function | | 1.4% | 100.0% | 100.0% |
| UGT1A1 | *6 | TPM/TPM2 | Genotype | Decreased Function | | 15.3% | 100.0% | 100.0% |
| | *27 | TPM/TPM2 | Genotype | Decreased Function | | 2.1% | 100.0% | 100.0% |
| | rs887829 | TPM/TPM2 | Genotype | Decreased Function | | 12.1% | 100.0% | 100.0% |
| VKORC1 | -1639G>A | TPM/TPM2 | Genotype | Require warfarin dose reduction | | 89.3% | 100.0% | 100.0% |

^ PharmGKB Gene-specific Information Tables https://www.pharmgkb.org/page/pgxGeneRef
PharmVar 5.0.1 https://www.pharmvar.org/
* PharmGKB Gene-specific Information Tables https://www.pharmgkb.org/page/pgxGeneRef
$ Evaluated the condordance of genotype and imputation calls with NGS or Sanger sequencing data

Table s2: Distribution of PGx phenotype, actionable phenotypes, and risk drug exposure

| Category | Gene | Phenotype | Actionable Phenotype | Case number | Phenotype Freq | | Risk drug | Risk drug exposure freq in each phenotype | Individuals encountered following risk drugs in each phenotype group | | | | | | | | | | | | | |
|---|---|---|---|---|---|---|---|---|---|---|---|---|---|---|---|---|---|---|---|---|---|---|
| | | | | | | | | | Efavirenz | Sertraline | | | | | | | | | | | | |
| Phase 1 Metabolic Enzymes | CYP2B6 | Total | | 486,956 | | | 13,118 | 2.7% | 258 | 4,786 | | | | | | | | | | | | |
| | | EM | | 318,276 | 65.4% | 318879 | 8,539 | 2.7% | 179 | 3,086 | | | | | | | | | | | | |
| | | IM | v | 149,998 | 30.8% | 168,077 | 4,082 | 2.7% | 73 | 1,507 | | | | | | | | | | | | |
| | | PM | v | 18,079 | 3.7% | | 487 | 2.7% | 6 | 189 | | | | | | | | | | | | |
| | | Indetermined | | 603 | 0.1% | | 10 | 1.7% | 0 | 4 | | | | | | | | | | | | |
| | | | | | | | | | Celecoxib | Flurbiprofen | Fluvastatin | Ibuprofen | Meloxicam | Nateglinide | Phenytoin | Piroxicam | Tenoxicam | Warfarin | | | | |
| | CYP2C9 | Total | | 486,956 | | | 92,103 | 18.9% | 50,200 | 28,989 | 4,219 | 6,681 | 10,174 | 227 | 670 | 13,933 | 23 | 6,113 | | | | |
| | | EM | | 451,020 | 92.6% | 451,393 | 85,301 | 18.9% | 46,570 | 26,807 | 3,903 | 6,135 | 9,447 | 210 | 623 | 12,841 | 23 | 5,665 | | | | |
| | | IM | v | 35,032 | 7.2% | 35,563 | 6,639 | 19.0% | 3,556 | 2,122 | 302 | 529 | 715 | 17 | 46 | 1,061 | - | 439 | | | | |
| | | PM | v | 531 | 0.1% | | 97 | 18.3% | 52 | 37 | 9 | 12 | 7 | | 1 | 17 | | 7 | | | | |
| | | Indetermined | | 373 | 0.1% | | 66 | 17.7% | 22 | 23 | 5 | 5 | 5 | | | 14 | | 2 | | | | |
| | | | | | | | | | Abrocitinib | Brivaracetam | Citalopram | Clobazam | Clopidogrel | Dexlansoprazol | Doxepin | Escitalopra | Imipramin | Lansoprazol | Omeprazole | Pantoprazol | Sertraline | Voriconazole |
| | | Total | | 486,956 | | | 116,120 | 23.8% | 11 | 28 | 6,942 | 1,283 | 33,664 | 19,147 | 6,773 | 6,845 | 9,636 | 44,378 | 33,407 | 28,138 | 4,786 | 343 |
| | CYP2C19 | UM | v | 17 | 0.0% | 187,877 | | 0.0% | | | | | | | | | | | | | | |
| | | RM | v | 3,183 | 0.7% | 299,079 | 751 | 23.6% | | | 51 | 6 | 229 | 122 | 44 | 51 | 46 | 284 | 210 | 185 | 30 | 3 |
| | | EM | | 187,095 | 38.4% | | 44,657 | 23.9% | 4 | 13 | 2,714 | 478 | 12,872 | 7,387 | 2,588 | 2,677 | 3,802 | 17,063 | 12,807 | 10,889 | 1,821 | 133 |
| | | IM | v | 226,301 | 46.5% | | 53,742 | 23.7% | 5 | 12 | 3,196 | 611 | 15,667 | 8,866 | 3,157 | 3,146 | 4,393 | 20,613 | 15,453 | 12,967 | 2,222 | 159 |
| | | PM | v | 69,578 | 14.3% | | 16,787 | 24.1% | 2 | 3 | 964 | 188 | 4,850 | 2,738 | 974 | 954 | 1,374 | 6,342 | 4,891 | 4,057 | 706 | 48 |
| | | na | | 782 | 0.2% | | 183 | 23.4% | | | 17 | | 46 | 34 | 10 | 17 | 21 | 76 | 46 | 40 | 7 | |
| | | | | | | | | | Tacrolimus | | | | | | | | | | | | | |
| | CYP3A5 | Total | | 486,956 | | | 3,299 | 0.7% | 3,299 | | | | | | | | | | | | | |
| | | PM | | 248,453 | 51.0% | 248,578 | 1,668 | 0.7% | 1,668 | | | | | | | | | | | | | |
| | | IM | v | 198,492 | 40.8% | 238,378 | 1,341 | 0.7% | 1,341 | | | | | | | | | | | | | |
| | | EM | v | 39,886 | 8.2% | | 289 | 0.7% | 289 | | | | | | | | | | | | | |
| | | Indetermined | | 125 | 0.0% | | 1 | 0.8% | 1 | | | | | | | | | | | | | |
| | | | | | | | | | Warfarin | | | | | | | | | | | | | |
| | CYP4F2 | Total | | 486,956 | | | 6,113 | 1.3% | 6,113 | | | | | | | | | | | | | |
| | | EM | | 284,173 | 58.4% | 285,017 | 3,578 | 1.3% | 3,578 | | | | | | | | | | | | | |
| | | IM | v | 174,577 | 35.8% | 201,939 | 2,174 | 1.2% | 2,174 | | | | | | | | | | | | | |
| | | PM | v | 27,362 | 5.6% | | 352 | 1.3% | 352 | | | | | | | | | | | | | |
| | | Indetermined | | 844 | 0.2% | | 9 | 1.1% | 9 | | | | | | | | | | | | | |
| | | | | | | | | | Isoniazid | Sulfamethoxazole and Trimethoprim | Sulfasalazine | | | | | | | | | | | |
| Phase 2 Metabolic Enzymes | NAT2 | Total | | 486,956 | | | 19,565 | 4.0% | 2,918 | 7,699 | 9,777 | | | | | | | | | | | |
| | | EM | | 139,510 | 28.6% | 382,014 | 5,597 | 4.0% | 854 | 2,172 | 2,803 | | | | | | | | | | | |
| | | IM | | 241,822 | 49.7% | 104,942 | 9,727 | 4.0% | 1,415 | 3,844 | 4,876 | | | | | | | | | | | |
| | | PM | v | 104,942 | 21.5% | | 4,205 | 4.0% | 646 | 1,666 | 2,082 | | | | | | | | | | | |
| | | Indetermined | | 682 | 0.1% | | 36 | 5.3% | 3 | 17 | 16 | | | | | | | | | | | |
| | | | | | | | | | Azathioprine | Mercaptopurine | | | | | | | | | | | | |
| | NUDT15 | Total | | 486,956 | | | 8,523 | 1.8% | 8,486 | 37 | | | | | | | | | | | | |
| | | EM | | 384,231 | 78.9% | 386,050 | 6,820 | 1.8% | 6,789 | 31 | | | | | | | | | | | | |
| | | IM | v | 95,046 | 19.5% | 100,906 | 1,595 | 1.7% | 1,590 | 5 | | | | | | | | | | | | |
| | | PM | v | 5,860 | 1.2% | | 70 | 1.2% | 70 | | | | | | | | | | | | | |
| | | Indetermined | | 1,819 | 0.4% | | 38 | 2.1% | 37 | 1 | | | | | | | | | | | | |
| | | | | | | | | | Azathioprine | Mercaptopurine | | | | | | | | | | | | |
| | TPMT | Total | | 486,956 | | | 8,523 | 1.8% | 8,486 | 37 | | | | | | | | | | | | |
| | | EM | | 473,144 | 97.2% | 473,360 | 8,279 | 1.7% | 8,243 | 36 | | | | | | | | | | | | |
| | | IM | v | 13,479 | 2.8% | 13,596 | 241 | 1.8% | 240 | 1 | | | | | | | | | | | | |
| | | PM | v | 117 | 0.0% | | 1 | 0.9% | 1 | | | | | | | | | | | | | |
| | | Indetermined | | 216 | 0.0% | | 2 | 0.9% | 2 | | | | | | | | | | | | | |
| | | | | | | | | | Atazanavir | Irinotecan | Nilotinib | Pazopanib | | | | | | | | | | |
| | UGT1A1 | Total | | 486,956 | | | 845 | 0.2% | 56 | 596 | 73 | 120 | | | | | | | | | | |
| | | EM | | 255,461 | 52.5% | 258,998 | 447 | 0.2% | 31 | 324 | 38 | 54 | | | | | | | | | | |
| | | IM | v | 176,543 | 36.3% | 227,958 | 304 | 0.2% | 19 | 208 | 28 | 49 | | | | | | | | | | |
| | | PM | v | 51,415 | 10.6% | | 84 | 0.2% | 6 | 56 | 6 | 16 | | | | | | | | | | |
| | | Indetermined | | 3,537 | 0.7% | | 10 | 0.3% | - | 8 | 1 | 1 | | | | | | | | | | |
| | | | | | | | | | Rosuvastatin | | | | | | | | | | | | | |
| Transproter | ABCG2 | Total | | 486,956 | | | 46,617 | 9.6% | 49,032 | | | | | | | | | | | | | |
| | | Normal | | 227,345 | 46.7% | 437,794 | 21,588 | 9.5% | 22,702 | | | | | | | | | | | | | |
| | | Decreased | | 210,293 | 43.2% | 49,162 | 20,314 | 9.7% | 21,384 | | | | | | | | | | | | | |
| | | Poor | v | 49,162 | 10.1% | | 4,703 | 9.6% | 4,932 | | | | | | | | | | | | | |
| | | Indetermined | | 156 | 0.0% | | 12 | 7.7% | 14 | | | | | | | | | | | | | |
| | | | | | | | | | Atorvastatin | Fluvastatin | Lovastatin | Pitavastatin | Pravastatin | Rosuvastatin | Simvastatin | | | | | | | |
| | SLCO1B1 | Total | | 486,956 | | | 127,197 | 26.1% | 73,078 | 4,219 | 4,154 | 32,334 | 7,786 | 49,032 | 11,558 | | | | | | | |
| | | Normal | | 383,660 | 78.8% | 383,711 | 100,390 | 26.2% | 57,718 | 3,314 | 3,248 | 25,416 | 6,140 | 38,557 | 9,000 | | | | | | | |
| | | Decreased | v | 97,031 | 19.9% | 103,245 | 25,163 | 25.9% | 14,440 | 847 | 839 | 6,498 | 1,540 | 9,817 | 2,391 | | | | | | | |
| | | Poor | v | 6,214 | 1.3% | | 1,637 | 26.3% | 915 | 58 | 67 | 419 | 106 | 655 | 166 | | | | | | | |
| | | Indetermined | | 51 | 0.0% | | 7 | 13.7% | 5 | - | - | 1 | - | 3 | 1 | | | | | | | |
| | | | | | | | | | Carbamazepine | | | | | | | | | | | | | |
| HLA | HLA-A*3101 | Total | | 486,956 | | | 1,319 | 0.3% | 1,319 | | | | | | | | | | | | | |
| | | Negative | | 467,890 | 96.1% | | 1,278 | 0.3% | 1,278 | | | | | | | | | | | | | |
| | | Positive | v | 19,066 | 3.9% | | 41 | 0.2% | 41 | | | | | | | | | | | | | |
| | | | | | | | | | Carbamazepin | Oxcarbazepine | Phenytoin | | | | | | | | | | | |
| | HLA-B*1502 | Total | | 486,956 | | | 3,961 | 0.8% | 1,319 | 2,229 | 670 | | | | | | | | | | | |

| | | | | | | | | | | | | | | |
|---|---|---|---|---|---|---|---|---|---|---|---|---|---|---|
| | | **Negative** | | 446,907 | 91.8% | | 3,675 | 0.8% | 1,247 | 2,056 | 616 | | | |
| | | **Positive** | **v** | 40,049 | 8.2% | | 286 | 0.7% | 72 | 173 | 54 | | | |
| | | | | | | | | | **Abacavir** | | | | | |
| | **HLA-B*5701** | **Total** | | 486,956 | | | 692 | 0.1% | 692 | | | | | |
| | | **Negative** | | 485,071 | 99.6% | | 691 | 0.1% | 691 | | | | | |
| | | **Positive** | **v** | 1,885 | 0.4% | | 1 | 0.1% | 1 | | | | | |
| | | | | | | | | | **Allopurinol** | | | | | |
| | **HLA-B*5801** | **Total** | | 486,956 | | | 5,251 | 1.1% | 5,251 | | | | | |
| | | **Negative** | | 389,717 | 80.0% | | 4,373 | 1.1% | 4,373 | | | | | |
| | | **Positive** | **v** | 97,239 | 20.0% | | 878 | 0.9% | 878 | | | | | |
| | | | | | | | | | **Lapatinib** | | | | | |
| | **DRB1*0701** | **Total** | | 486,956 | | | 27 | 0.0% | 27 | | | | | |
| | | **Negative** | | 464,494 | 95.4% | | 27 | 0.0% | 27 | | | | | |
| | | **Positive** | **v** | 22,462 | 4.6% | | | 0.0% | | | | | | |
| | | | | | | | | | **Lapatinib** | | | | | |
| | **DQA1*0201** | **Total** | | 486,956 | | | 27 | 0.0% | 27 | | | | | |
| | | **Negative** | | 464,545 | 95.4% | | 27 | 0.0% | 27 | | | | | |
| | | **Positive** | **v** | 22,411 | 4.6% | | | 0.0% | | | | | | |
| | | | | | | | | | **Dapsone** | **Nitrofurantoin** | **Rasburicase** | | | |
| **Others** | **G6PD** | **Total** | | 486,956 | | | 144 | 0.0% | 108 | 5 | 31 | | | |
| | | **normal** | | 446,940 | 91.8% | 466,603 | 129 | 0.0% | 96 | 4 | 29 | | | |
| | | **variable** | **v** | 14,095 | 2.9% | 20,353 | 6 | 0.0% | 5 | 1 | | | | |
| | | **deficient** | **v** | 6,258 | 1.3% | | 3 | 0.0% | 2 | | 1 | | | |
| | | **Indetermined** | | 19,663 | 4.0% | | 6 | 0.0% | 5 | | 1 | | | |
| | | | | | | | | | **Warfarin** | | | | | |
| | **VKORC1** | **Total** | | 486,956 | | | 6,113 | 1.3% | 6,113 | | | | | |
| | | **0** | **v** | 388,233 | 79.7% | 5,796 | 4,803 | 1.2% | 4,803 | | | | | |
| | | **1** | **v** | 92,927 | 19.1% | 481,160 | 1,237 | 1.3% | 1,237 | | | | | |
| | | **2** | | 5,676 | 1.2% | | 70 | 1.2% | 70 | | | | | |
| | | **Indetermined** | | 120 | 0.0% | | 3 | 2.5% | 3 | | | | | |
| | | | | | | | | | **Peginterferon alfa-2a** | **Peginterferon alfa-2b** | **Ribavirin** | | | |
| | **IFNL3** | **Total** | | 486,956 | | | 4,255 | 0.9% | 576 | 218 | 1,873 | | | |
| | | **Favored** | | 431,219 | 88.6% | 431,571 | 3,692 | 0.9% | 504 | 181 | 1,591 | | | |
| | | **Unfavored** | **v** | 55,385 | 11.4% | 55,385 | 562 | 1.0% | 72 | 37 | 282 | | | |
| | | **Indetermined** | | 352 | 0.1% | | 1 | 0.3% | | | | | | |
| | | | | | | | | | **Amikacin** | **Gentamicin** | **Kanamycin** | **Paromomycin** | **Streptomycin** | **Tobramycin** |
| | **MT-RNR1** | **Total** | | 486,956 | | | 27,595 | 5.7% | 635 | 23,953 | 5 | 15 | 59 | 4,190 |
| | | **Normal** | | 484,521 | 99.5% | 485,059 | 27,441 | 5.7% | 631 | 23,816 | 5 | 15 | 59 | 4,168 |
| | | **Risk** | **v** | 1,897 | 0.4% | 1,897 | 121 | 6.4% | 3 | 109 | | | | 16 |
| | | **Indetermined** | | 538 | 0.1% | | 33 | 6.1% | 1 | 28 | | | | 6 |

**Table s3: The distribution of TPMI participants exposed to high-risk PGx drugs**

| Number of drug prescibed per person | Case number | Accumulated count | Accumulated % |
|---|---|---|---|
| 0 | 249796 | | |
| 1 | 98724 | 237160 | 48.7% |
| 2 | 60111 | 138436 | 28.4% |
| 3 | 33926 | 78325 | 16.1% |
| 4 | 19251 | 44399 | 9.1% |
| 5 | 10596 | 25148 | 5.2% |
| 6 | 6114 | 14552 | 3.0% |
| 7 | 3550 | 8438 | 1.7% |
| 8 | 2124 | 4888 | 1.0% |
| 9 | 1133 | 2764 | 0.6% |
| 10 | 673 | 1631 | 0.3% |
| 11 | 440 | 958 | 0.2% |
| 12 | 242 | 518 | 0.1% |
| 13 | 132 | 276 | 0.1% |
| 14 | 78 | 144 | 0.0% |
| 15 | 42 | 66 | 0.0% |
| 16 | 14 | 24 | 0.0% |
| 17 | 9 | 10 | 0.0% |
| 18 | 1 | 1 | 0.0% |

**Table s4: The demography of clopidogrel-related MACE cohort**

| Clopidogrel Users | no MACE | | MACE | | OR (95% CI) | P value |
|---|---|---|---|---|---|---|
| No. of subjects | 24578 | | 3477 | | | |
| Male | 8208 | 33.4% | 1179 | 33.9% | 0.98 (0.91-1.06) | 5.49E-01 |
| Age (years, mean±SD) | 72.0±9.5 | | 71.24±10.4 | | | 1.50E-01 |
| Comorbidity | | | | | | |
| DM | 13700 | 55.7% | 1910 | 54.9% | 0.97 (0.9-1.04) | 3.7E-01 |
| HC | 15803 | 64.3% | 2219 | 63.8% | 0.98 (0.91-1.06) | 5.8E-01 |
| HTN | 11503 | 46.8% | 1625 | 46.7% | 1 (0.93-1.07) | 9.1E-01 |
| CKD | 8852 | 36.0% | 1209 | 34.8% | 0.95 (0.88-1.02) | 1.5E-01 |
| Concurrent medications | | | | | | |
| aspirin | 2231 | 9.1% | 324 | 9.3% | 1.03 (0.91-1.16) | 6.4E-01 |
| warfarin | 1253 | 5.1% | 190 | 5.5% | 1.08 (0.92-1.26) | 3.6E-01 |
| PPI | 8670 | 35.3% | 1420 | 40.8% | 1.27 (1.18-1.37) | 1.56E-10 |
| statin | 10061 | 40.9% | 1458 | 41.9% | 1.04 (0.97-1.12) | 2.8E-01 |

**Table s5: Influence of CYP2C19 for incidence of MACE**

**1. Clopidogrel users with one or two CYP2C19 LoF vs with non-LoF**

| | Univariate analysis | | | | Multivariate analysis | | | |
|---|---|---|---|---|---|---|---|---|
| MACE | OR | Lower95%CI | Upper95%CI | Pvalue | OR | Lower95%CI | Upper95%CI | Pvalue |
| Stroke | 1.007 | 1.004 | 1.011 | 7.30E-05 | 1.007 | 1.004 | 1.011 | 7.10E-05 |
| MI | 1.023 | 1.019 | 1.026 | 8.30E-37 | 1.023 | 1.019 | 1.026 | 2.56E-37 |
| UA | 1.014 | 1.010 | 1.018 | 2.21E-13 | 1.014 | 1.010 | 1.018 | 1.61E-13 |
| HF | 1.014 | 1.009 | 1.019 | 1.31E-07 | 1.014 | 1.009 | 1.019 | 8.20E-08 |
| TLR | 1.009 | 1.004 | 1.013 | 1.28E-04 | 1.009 | 1.004 | 1.014 | 1.16E-04 |
| CV death | 1.001 | 0.999 | 1.002 | 2.35E-01 | 1.001 | 0.999 | 1.002 | 2.36E-01 |
| MACE | 1.045 | 1.036 | 1.053 | 2.10E-27 | 1.045 | 1.037 | 1.053 | 7.98E-28 |

**2. Clopidogrel users with one CYP2C19 LoF vs with non-LoF**

| | Univariate analysis | | | | Multivariate analysis | | | |
|---|---|---|---|---|---|---|---|---|
| MACE | OR | Lower95%CI | Upper95%CI | Pvalue | OR | Lower95%CI | Upper95%CI | Pvalue |
| Stroke | 1.007 | 1.003 | 1.011 | 2.77E-04 | 1.00706 | 1.00326 | 1.01088 | 2.69E-04 |
| MI | 1.022 | 1.019 | 1.026 | 1.83E-34 | 1.02242 | 1.01883 | 1.02602 | 6.11E-35 |
| UA | 1.013 | 1.010 | 1.017 | 1.40E-11 | 1.01355 | 1.00963 | 1.01748 | 1.03E-11 |
| HF | 1.012 | 1.007 | 1.018 | 5.00E-06 | 1.01260 | 1.00727 | 1.01796 | 3.37E-06 |
| TLR | 1.008 | 1.004 | 1.013 | 5.76E-04 | 1.00839 | 1.00363 | 1.01318 | 5.45E-04 |
| CV death | 1.001 | 0.999 | 1.002 | 3.19E-01 | 1.00077 | 0.99924 | 1.00230 | 3.22E-01 |
| MACE | 1.042 | 1.034 | 1.051 | 6.97E-23 | 1.04262 | 1.03407 | 1.05125 | 3.28E-23 |

**3. Clopidogrel users with two CYP2C19 LoF vs with non-LoF**

| | Univariate analysis | | | | Multivariate analysis | | | |
|---|---|---|---|---|---|---|---|---|
| MACE | OR | Lower95%CI | Upper95%CI | Pvalue | OR | Lower95%CI | Upper95%CI | Pvalue |
| Stroke | 1.008 | 1.003 | 1.014 | 1.62E-03 | 1.008 | 1.003 | 1.014 | 1.62E-03 |
| MI | 1.024 | 1.020 | 1.028 | 6.71E-28 | 1.024 | 1.020 | 1.028 | 4.27E-28 |
| UA | 1.016 | 1.011 | 1.022 | 6.04E-10 | 1.017 | 1.011 | 1.022 | 5.45E-10 |
| HF | 1.018 | 1.011 | 1.026 | 1.00E-06 | 1.018 | 1.011 | 1.026 | 1.00E-06 |
| TLR | 1.011 | 1.004 | 1.017 | 1.65E-03 | 1.011 | 1.004 | 1.017 | 1.58E-03 |
| CV death | 1.001 | 0.999 | 1.003 | 2.54E-01 | 1.001 | 0.999 | 1.003 | 2.56E-01 |
| MACE | 1.053 | 1.041 | 1.064 | 8.38E-19 | 1.053 | 1.041 | 1.065 | 5.74E-19 |

**Table s6: Patient characteristics of azathioprine users in TPMI**

| Azathioprine Users | no ADR | | ADR | | OR (95% CI) | P value |
|---|---|---|---|---|---|---|
| No. of subjects | 6948 | | 1503 | | | |
| Male | 2140 | 30.8% | 434 | 28.9% | 0.91 (0.8-1.03) | 1.40E-01 |
| Age (years, mean±SD) | 46.2±15 | | 42.5±14.7 | | | 9.43E-02 |
| Comorbidity | | | | | | |
| DM | 763 | 11.0% | 170 | 11.3% | 1.03 (0.86-1.23) | 7.10E-01 |
| HC | 1329 | 19.1% | 305 | 20.3% | 1.08 (0.94-1.24) | 3.00E-01 |
| HTN | 1304 | 18.8% | 309 | 20.6% | 1.12 (0.97-1.37) | 1.10E-01 |
| IHD | 484 | 7.0% | 115 | 7.7% | 1.11 (0.9-1.37) | 3.50E-01 |
| HF | 138 | 2.0% | 34 | 2.3% | 1.14 (0.76-1.67) | 4.90E-01 |
| CVD | 292 | 4.2% | 72 | 4.8% | 1.15 (0.88-1.5) | 3.10E-01 |
| Concurrent medications | | | | | | |
| aspirin | 327 | 4.7% | 61 | 4.1% | 0.86 (0.65-1.14) | 2.80E-01 |
| allopurinol | 67 | 1.0% | 29 | 1.9% | 2.02 (1.3-3.13) | 1.37E-03 |
| corticosteroids | 5262 | 75.7% | 1164 | 77.4% | 1.1 (0.96-1.26) | 1.60E-01 |
| methotrexate | 1030 | 14.8% | 217 | 14.4% | 0.97 (0.83-1.14) | 7.00E-01 |

**Table s7: Influence of NUDT15 and TPMT for azathioprine discontinuation due to ADR**

**1. Azathioprine users with one or two NUDT15 LoF vs with non-LoF**

| | Univariate analysis | | | | Multivariate analysis | | | |
|---|---|---|---|---|---|---|---|---|
| ADR | OR | Lower95%CI | Upper95%C | Pvalue | OR | Lower95%CI | Upper95%C | Pvalue |
| Leukopenia | 1.0669 | 1.0494 | 1.0848 | 0.0000 | 1.0668 | 1.0493 | 1.0847 | 2.14E-14 |
| Thrombocytopen | 1.0103 | 0.9967 | 1.0241 | 0.1390 | 1.0117 | 0.9981 | 1.0255 | 9.17E-02 |
| Hepatitis | 0.9991 | 0.9879 | 1.0105 | 0.8820 | 0.9994 | 0.9883 | 1.0107 | 9.23E-01 |
| Allergy | 1.0009 | 0.9968 | 1.0050 | 0.6710 | 1.0003 | 0.9961 | 1.0046 | 8.77E-01 |
| GI | 1.0028 | 0.9973 | 1.0083 | 0.3140 | 1.0037 | 0.9982 | 1.0092 | 1.89E-01 |
| HairLoss | 1.0014 | 0.9970 | 1.0058 | 0.5440 | 1.0010 | 0.9966 | 1.0055 | 6.42E-01 |
| ADR | 1.0461 | 1.0248 | 1.0679 | 0.0000 | 1.0478 | 1.0264 | 1.0696 | 9.28E-06 |

**2. Azathioprine users with one NUDT15 LoF vs with non-LoF**

| | Univariate analysis | | | | Multivariate analysis | | | |
|---|---|---|---|---|---|---|---|---|
| ADR | OR | Lower95%CI | Upper95%C | Pvalue | OR | Lower95%CI | Upper95%C | Pvalue |
| Leukopenia | 1.0568 | 1.0393 | 1.0746 | 0.0000 | 1.0574 | 1.0399 | 1.0752 | 6.33E-11 |
| Thrombocytopen | 1.0066 | 0.9929 | 1.0205 | 0.3459 | 1.0084 | 0.9947 | 1.0223 | 2.30E-01 |
| Hepatitis | 0.9980 | 0.9691 | 1.0227 | 0.7426 | 0.9981 | 0.9868 | 1.0096 | 7.44E-01 |
| Allergy | 1.0006 | 0.9964 | 1.0047 | 0.7950 | 0.9999 | 0.9957 | 1.0042 | 9.78E-01 |
| GI | 1.0021 | 0.9966 | 1.0077 | 0.4559 | 1.0030 | 0.9974 | 1.0086 | 2.94E-01 |
| HairLoss | 0.9986 | 0.9943 | 1.0029 | 0.5097 | 0.9988 | 0.9944 | 1.0031 | 5.75E-01 |
| ADR | 1.0348 | 1.0135 | 1.0566 | 0.0013 | 1.0376 | 1.0161 | 1.0595 | 5.39E-04 |

**3. Azathioprine users with two NUDT15 LoF vs with non-LoF**

| | Univariate analysis | | | | Multivariate analysis | | | |
|---|---|---|---|---|---|---|---|---|
| ADR | OR | Lower95%CI | Upper95%C | Pvalue | OR | Lower95%CI | Upper95%C | Pvalue |
| Leukopenia | 1.327 | 1.238 | 1.423 | 2.26E-15 | 1.312 | 1.223 | 1.408 | 3.55E-14 |
| Thrombocytopen | 1.098 | 1.035 | 1.165 | 2.04E-03 | 1.091 | 1.028 | 1.158 | 4.04E-03 |
| Hepatitis | 1.027 | 0.976 | 1.079 | 3.04E-01 | 1.032 | 0.981 | 1.085 | 2.24E-01 |
| Allergy | 1.009 | 0.991 | 1.027 | 3.45E-01 | 1.010 | 0.991 | 1.029 | 3.19E-01 |
| GI | 1.019 | 0.995 | 1.044 | 1.15E-01 | 1.021 | 0.997 | 1.045 | 9.37E-02 |
| HairLoss | 1.068 | 1.047 | 1.090 | 9.89E-11 | 1.056 | 1.035 | 1.077 | 1.05E-07 |
| ADR | 1.342 | 1.228 | 1.468 | 1.09E-10 | 1.316 | 1.203 | 1.440 | 2.33E-09 |

**4. Azathioprine users with one or two TPMT LoF vs with non-LoF**

| | Univariate analysis | | | | Multivariate analysis | | | |
|---|---|---|---|---|---|---|---|---|
| ADR | OR | Lower95%CI | Upper95%C | Pvalue | OR | Lower95%CI | Upper95%C | Pvalue |
| Leukopenia | 1.006 | 0.967 | 1.046 | 0.778 | 1.003 | 0.964 | 1.043 | 8.96E-01 |
| Thrombocytopen | 0.994 | 0.963 | 1.027 | 0.727 | 0.999 | 0.967 | 1.031 | 9.39E-01 |
| Hepatitis | 0.995 | 0.969 | 1.022 | 0.733 | 0.994 | 0.968 | 1.021 | 6.69E-01 |
| Allergy | 0.998 | 0.988 | 1.008 | 0.717 | 0.998 | 0.988 | 1.008 | 7.21E-01 |
| GI | 0.998 | 0.985 | 1.011 | 0.743 | 0.998 | 0.985 | 1.012 | 8.09E-01 |
| HairLoss | 1.006 | 0.995 | 1.016 | 0.273 | 1.002 | 0.991 | 1.012 | 7.35E-01 |
| ADR | 0.988 | 0.941 | 1.037 | 0.624 | 0.988 | 0.941 | 1.038 | 6.40E-01 |

**5. Azathioprine users with one TPMT LoF vs with non-LoF**

| | Univariate analysis | | | | Multivariate analysis | | | |
|---|---|---|---|---|---|---|---|---|
| ADR | OR | Lower95%CI | Upper95%C | Pvalue | OR | Lower95%CI | Upper95%C | Pvalue |
| Leukopenia | 1.006 | 0.967 | 1.047 | 0.761 | 1.003 | 0.964 | 1.044 | 8.76E-01 |
| Thrombocytopen | 0.995 | 0.963 | 1.027 | 0.739 | 0.999 | 0.967 | 1.032 | 9.47E-01 |
| Hepatitis | 0.996 | 0.969 | 1.023 | 0.743 | 0.994 | 0.968 | 1.021 | 6.77E-01 |
| Allergy | 0.998 | 0.988 | 1.008 | 0.720 | 0.998 | 0.988 | 1.008 | 7.24E-01 |
| GI | 0.998 | 0.985 | 1.011 | 0.747 | 0.998 | 0.985 | 1.012 | 8.13E-01 |
| HairLoss | 1.006 | 0.995 | 1.017 | 0.269 | 1.002 | 0.991 | 1.012 | 7.28E-01 |
| ADR | 0.988 | 0.941 | 1.038 | 0.644 | 0.989 | 0.942 | 1.039 | 6.59E-01 |

**Table s8: Influence of ABCG2, CYP2C9 and SLCO1B1 for statin-associated myopathy**

**1. Atorvastatin users with decreased or poor function ABCG2 vs with normal function ABCG2**

| | ADR(non LOF) | ADR(LOF) | no ADR (non LOF) | no ADR (LOF) | OR | Upper 95%CI | Lower 95%CI | p value |
|---|---|---|---|---|---|---|---|---|
| HighCPKonly | 2 | 1 | 33787 | 39264 | 0.43 | 4.74 | 0.04 | 4.78E-01 |
| myalgia | 368 | 466 | 33421 | 38799 | 1.09 | 1.25 | 0.95 | 2.15E-01 |
| myositis | 38 | 52 | 33751 | 39213 | 1.18 | 1.79 | 0.78 | 4.43E-01 |
| rhabdomyolysis | 7 | 15 | 33782 | 39250 | 1.84 | 4.51 | 0.75 | 1.74E-01 |
| SAM | 415 | 534 | 33374 | 38731 | 1.11 | 1.26 | 0.98 | 1.17E-01 |
| sSAM | 45 | 67 | 33744 | 39198 | 1.28 | 1.87 | 0.88 | 1.97E-01 |

**2. Atorvastatin users with decreased function ABCG2 vs with normal function ABCG2**

| | ADR(non LOF) | ADR(LOF) | no ADR (non LOF) | no ADR (LOF) | OR | Upper 95%CI | Lower 95%CI | p value |
|---|---|---|---|---|---|---|---|---|
| HighCPKonly | 2 | 1 | 33787 | 31738 | 0.53 | 5.85 | 0.05 | 5.36E-01 |
| myalgia | 368 | 375 | 33421 | 31364 | 1.09 | 1.26 | 0.94 | 2.25E-01 |
| myositis | 38 | 33 | 33751 | 31706 | 0.92 | 1.47 | 0.58 | 5.19E-01 |
| rhabdomyolysis | 7 | 15 | 33782 | 31724 | 2.28 | 5.59 | 0.93 | 1.03E-01 |
| SAM | 415 | 424 | 33374 | 31315 | 1.09 | 1.25 | 0.95 | 1.47E-01 |
| sSAM | 45 | 48 | 33744 | 31691 | 1.14 | 1.71 | 0.76 | 2.82E-01 |

**3. Atorvastatin users with poor function ABCG2 vs with normal function ABCG2**

| | ADR(non LOF) | ADR(LOF) | no ADR (non LOF) | no ADR (LOF) | OR | Upper 95%CI | Lower 95%CI | p value |
|---|---|---|---|---|---|---|---|---|
| HighCPKonly | 2 | 0 | 33787 | 7526 | | | | 5.04E-01 |
| myalgia | 368 | 91 | 33421 | 7435 | 1.11 | 1.4 | 0.88 | 3.69E-01 |
| myositis | 38 | 19 | 33751 | 7507 | 2.25 | 3.9 | 1.3 | 3.09E-03 |
| rhabdomyolysis | 7 | 0 | 33782 | 7526 | | | | 2.12E-01 |
| SAM | 415 | 110 | 33374 | 7416 | 1.19 | 1.47 | 0.96 | 1.02E-01 |
| sSAM | 45 | 19 | 33744 | 7507 | 1.9 | 3.25 | 1.11 | 1.73E-02 |

**6. Atorvastatin users with decreased or poor function SLCO1B1 vs with normal function SLCO1B1**

| | ADR(non LOF) | ADR(LOF) | no ADR (non LOF) | no ADR (LOF) | OR | Upper 95%CI | Lower 95%CI | p value |
|---|---|---|---|---|---|---|---|---|
| HighCPKonly | 2 | 1 | 57716 | 15354 | 1.88 | 20.74 | 0.17 | 6.00E-01 |
| myalgia | 625 | 209 | 57093 | 15146 | 1.26 | 1.48 | 1.08 | 3.91E-03 |
| myositis | 67 | 23 | 57651 | 15332 | 1.29 | 2.07 | 0.8 | 2.90E-01 |
| rhabdomyolysis | 17 | 5 | 57701 | 15350 | 1.11 | 3.01 | 0.41 | 8.44E-01 |
| SAM | 711 | 238 | 57007 | 15117 | 1.26 | 1.46 | 1.09 | 1.97E-03 |
| sSAM | 84 | 28 | 57634 | 15327 | 1.25 | 1.92 | 0.81 | 3.00E-01 |

**7. Atorvastatin users with decreased function SLCO1B1 vs with normal function SLCO1B1**

| | ADR(non LOF) | ADR(LOF) | no ADR (non LOF) | no ADR (LOF) | OR | Upper 95%CI | Lower 95%CI | p value |
|---|---|---|---|---|---|---|---|---|
| HighCPKonly | 2 | 1 | 57716 | 14439 | 2 | 22.06 | 0.18 | 5.70E-01 |
| myalgia | 625 | 190 | 57093 | 14250 | 1.22 | 1.44 | 1.04 | 1.24E-02 |
| myositis | 67 | 22 | 57651 | 14418 | 1.31 | 2.12 | 0.81 | 2.69E-01 |
| rhabdomyolysis | 17 | 5 | 57701 | 14435 | 1.18 | 3.2 | 0.44 | 7.65E-01 |
| SAM | 711 | 218 | 57007 | 14222 | 1.23 | 1.43 | 1.06 | 5.73E-03 |
| sSAM | 84 | 27 | 57634 | 14413 | 1.29 | 1.99 | 0.84 | 2.62E-01 |

**8. Atorvastatin users with poor function SLCO1B1 vs with normal function SLCO1B1**

| | ADR(non LOF) | ADR(LOF) | no ADR (non LOF) | no ADR (LOF) | OR | Upper 95%CI | Lower 95%CI | p value |
|---|---|---|---|---|---|---|---|---|
| HighCPKonly | 2 | 0 | 57716 | 915 | | | | |
| myalgia | 625 | 19 | 57093 | 896 | 1.94 | 3.08 | 1.22 | 4.22E-03 |
| myositis | 67 | 1 | 57651 | 914 | 0.94 | 6.78 | 0.13 | 9.52E-01 |
| rhabdomyolysis | 17 | 0 | 57701 | 915 | | | | |
| SAM | 711 | 20 | 57007 | 895 | 1.79 | 2.81 | 1.14 | 9.87E-03 |
| sSAM | 84 | 1 | 57634 | 914 | 0.75 | 5.39 | 0.1 | 7.75E-01 |

**Table s9: Patient characteristics of NSAID users in TPMI**

| NSAID Users | no ADR | | ADR | | OR (95% CI) | P value |
|---|---|---|---|---|---|---|
| No. of subjects | 9471 | | 529 | | | |
| Male | 3288 | 34.7% | 205 | 38.8% | 1.19 (0.99-1.42) | 6.00E-02 |
| Age (years, mean±SD) | 61±14 | | 61.8±14.3 | | | 1.10E-01 |
| Dose_first (DDD) | 1.1±0.84 | | 1.22±0.4 | | | 6.00E-02 |
| Comorbidity | | | | | | |
| HTN | 1857 | 19.6% | 213 | 40.3% | 2.76 (2.3-3.31) | 3.62E-30 |
| HC | 1940 | 20.5% | 178 | 33.6% | 1.97 (1.63-2.38) | 5.51E-13 |
| DM | 1466 | 15.5% | 159 | 30.1% | 2.35 (1.94-2.85) | 9.15E-19 |
| CVD | 500 | 5.3% | 65 | 12.3% | 2.51 (1.91-3.3) | 1.09E-11 |
| IHD | 817 | 8.6% | 80 | 15.1% | 1.89 (1.47-2.42) | 3.60E-07 |
| PVD | 23 | 0.2% | 11 | 2.1% | 8.72 (4.23-17.99) | 1.64E-12 |
| COPD | 43 | 0.5% | 14 | 2.6% | 5.96 (3.24-10.96) | 7.09E-11 |
| HF | 122 | 1.3% | 35 | 6.6% | 5.43 (3.69-7.99) | 8.50E-22 |
| HP | 6 | 0.1% | 0 | 0.0% | | 5.63E-01 |
| Concurrent medications | | | | | | |
| aspirin | 769 | 8.1% | 88 | 16.6% | 2.26 (1.78-2.87) | 9.80E-12 |
| antibiotics | 3 | 0.0% | 5 | 0.9% | 30.11 (7.18-126.34) | 4.76E-13 |
| ACEI/ARB | 20 | 0.2% | 9 | 1.7% | 8.18 (3.71-18.05) | 5.55E-10 |
| diuretics | 66 | 0.7% | 80 | 15.1% | 25.39 (18.08-35.65) | 1.25E-159 |
| anticoagulants | 45 | 0.5% | 5 | 0.9% | 2 (0.79-5.06) | 1.36E-01 |
| immunosuppressants | 36 | 0.4% | 11 | 2.1% | 5.57 (2.82-11.01) | 2.68E-08 |
| CYP2C9 | | | | | | |
| EM | 8757 | 92.5% | 499 | 94.3% | | |
| IM | 693 | 7.3% | 28 | 5.3% | 0.71 (0.48-1.05) | 8.13E-02 |
| PM | 12 | 0.1% | 2 | 0.4% | 2.92 (0.65-13.08) | 1.41E-01 |
| Indetermined | 9 | 0.1% | 0 | 0.0% | | 4.74E-01 |

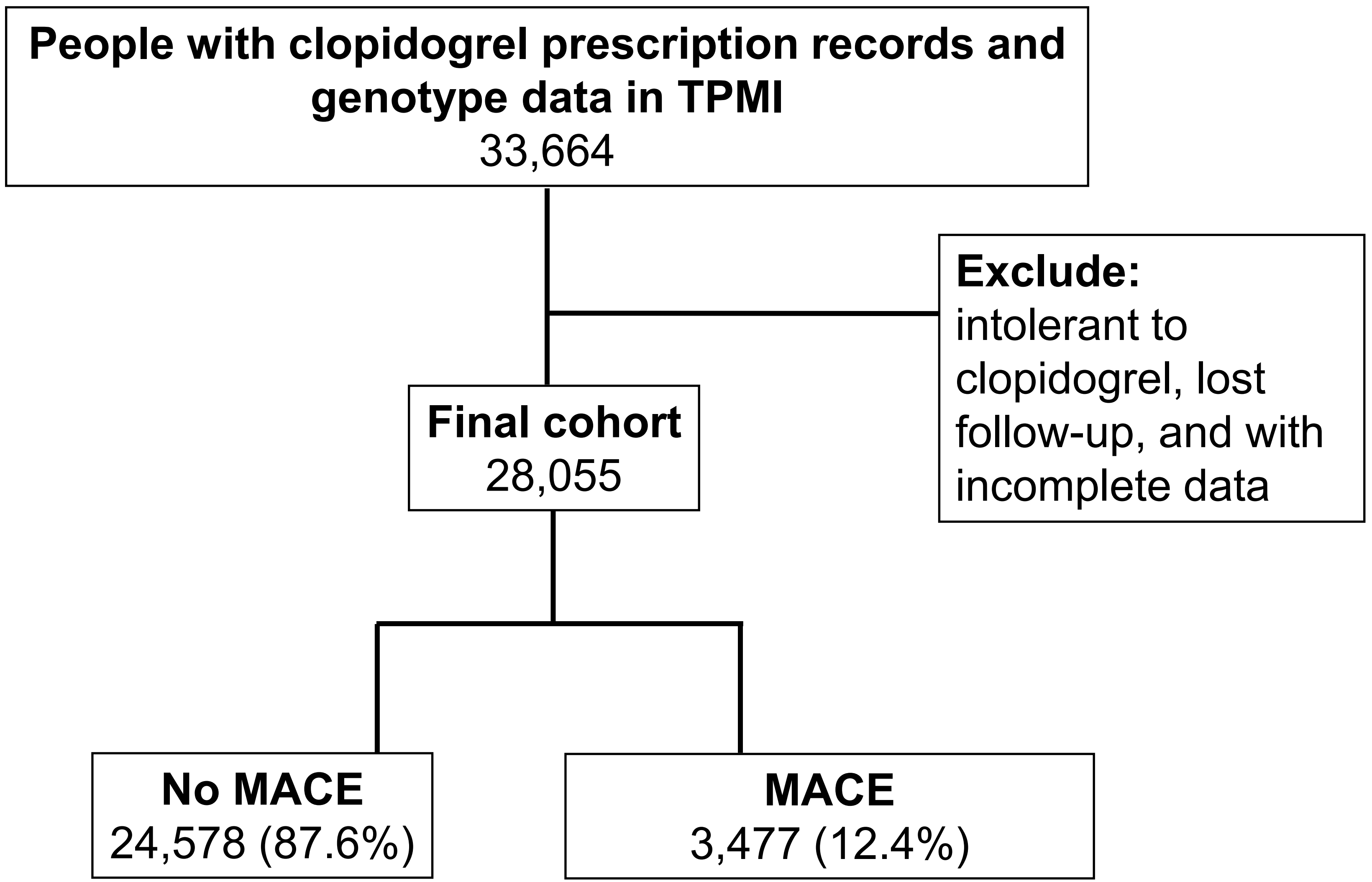


**Figure s1: study design for clopidogrel-related adverse drug reactions**

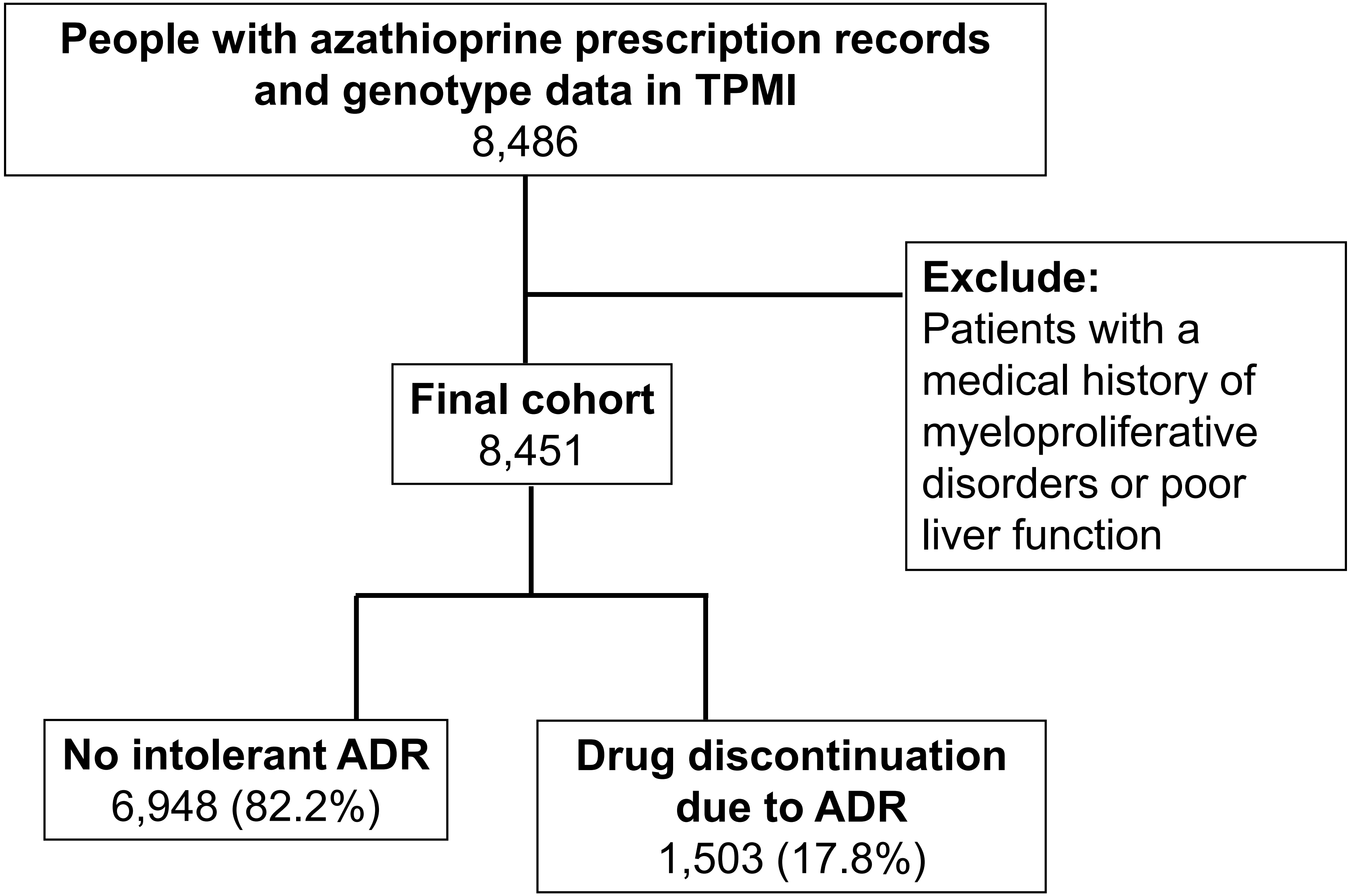


**Figure s2: study design for azathioprine-related adverse drug reactions**
